# Disease stage-specific atrophy markers in Alzheimer’s disease

**DOI:** 10.1101/2025.03.13.25323904

**Authors:** Hannah Baumeister, Helena M. Gellersen, Sarah E. Polk, René Lattmann, Anika Wuestefeld, Laura E. M. Wisse, Trevor Glenn, Renat Yakupov, Melina Stark, Luca Kleineidam, Sandra Roeske, Barbara Marcos Morgado, Hermann Esselmann, Frederic Brosseron, Alfredo Ramirez, Falk Lüsebrink, Matthis Synofzik, Björn H. Schott, Matthias C. Schmid, Stefan Hetzer, Peter Dechent, Klaus Scheffler, Michael Ewers, Julian Hellmann-Regen, Ersin Ersözlü, Eike Spruth, Maria Gemenetzi, Klaus Fliessbach, Claudia Bartels, Ayda Rostamzadeh, Wenzel Glanz, Enise I. Incesoy, Daniel Janowitz, Boris-Stephan Rauchmann, Ingo Kilimann, Sebastian Sodenkamp, Marie Coenjaerts, Annika Spottke, Oliver Peters, Josef Priller, Anja Schneider, Jens Wiltfang, Katharina Buerger, Robert Perneczky, Stefan Teipel, Christoph Laske, Michael Wagner, Gabriel Ziegler, Frank Jessen, Emrah Düzel, David Berron, the DELCODE study group

## Abstract

**INTRODUCTION:** Structural MRI often lacks diagnostic, prognostic, and monitoring value in Alzheimer’s disease (AD), particularly in early disease stages. To improve its utility, we aimed to identify optimal MRI readouts for different use cases.

**METHODS:** We included 363 older adults; healthy controls (HC) who were negative or positive for amyloid-beta (Aβ) and Aβ-positive patients with subjective cognitive decline (SCD), mild cognitive impairment, or dementia of the Alzheimer type. MRI and neuropsychological assessments were administered annually for up to three years.

**RESULTS:** Accelerated atrophy of distinct MTL subregions was evident already during preclinical AD. Symptomatic disease stages most notably differed in their hippocampal and parietal atrophy signatures. Associations of atrophy markers and cognitive inventories varied by intended use and disease stage.

**DISCUSSION:** With the appropriate readout, MRI can detect abnormal atrophy already during preclinical AD. To optimize performance, MRI readouts should be tailored to the targeted disease stage and intended use.

## Background

Aside from accumulations of amyloid-beta (Aβ) and neurofibrillary tangles (NFTs) of hyperphosphorylated tau, neurodegeneration is a core pathological hallmark of Alzheimer’s disease (AD).^1^ Structural MRI is commonly used for assessing atrophy as a downstream biomarker of neurodegeneration, offering advantages like non-invasiveness, good accessibility, and robust associations with clinical phenotypes.^2,3^ However, its current routine use lacks sensitivity especially to early disease stages—a crucial property as evidence highlights preclinical and prodromal disease stages as the optimal window for disease-modifying treatment.^4,5^

A biomarker’s performance may not just vary by disease stage, but also by use case. Biomarker uses are manifold and include clinical diagnosis and prognosis as well as disease monitoring. In pharmacological trials, biomarkers can aid in aspects like case finding, sample enrichment or stratification, as well as evaluating treatment efficacy and safety.^1,5,6^ Thus, establishing sensitive MRI readouts for various contexts-of-use and disease stages requires comprehensive knowledge of each candidate’s characteristics, such as earliest abnormality, rates of change along the disease course, and relationships with neuropsychological inventories.^1^

### Candidate readouts for improving MRI performance

With prior studies demonstrating a co-localization of NFTs and subsequent hotspots of neurodegeneration, MRI sensitivity may be improved by anchoring readout selection to the (presumed) NFT accumulation stage of interest.^3,7–9^ The medial temporal lobe (MTL) is among the earliest brain regions affected by AD-related NFTs and its subregions—including hippocampal subfields, the amygdala, and cortical structures—are thought to exhibit varying degrees and timelines of NFT accumulation.^8,10–13^ This makes MTL subregional volumes promising atrophy readouts for providing fine-grained insights into disease progression in early AD, such as in patients with minimal or no cognitive impairment or in preclinical AD intervention trials. As the disease advances, NFTs increasingly occur in the parietal lobe.^8,14^ Not only do MTL and parietal structures exhibit AD pathology at different disease stages, but they also serve highly specialized functional roles in the memory system.^15^ Thus, atrophy in these regions may not only reflect biological disease progression, but may also be associated with the deterioration of specific cognitive functions. Though it has been shown that MTL and parietal atrophy are related to cognitive decline, the potentially dynamic coupling of subregional MRI readouts and established neuropsychological inventories across the AD continuum are needed to inform marker selection for different applications.^16^

### Challenges in longitudinal MRI assessments

Longitudinal assessments of brain structure show promise for early atrophy detection. With each participant serving as their own reference, abnormal atrophy rates can be detected before absolute readouts fall below a population norm. Additionally, the influence of inter-individual anatomical variations is mitigated, which is particularly relevant within the MTL where high anatomical variability may overshadow subtle disease-related effects.^11,17^

When performing longitudinal segmentations, processing images individually may introduce unrelated noise. Hence, pipelines that minimize and homogenize noise across measurements by using within-subject templates are increasingly popular.^18^

### The current study

While there have been numerous studies on AD-related atrophy, no comprehensive research has examined the disease stage-specific properties of both cross-sectionally and longitudinally recorded subregional MTL and parietal atrophy markers for different use cases.^19–27^ Additionally, many studies were limited by lacking biomarker evidence of AD pathology and a subsequent inability to identify preclinical cases. Finally, highly controlled settings often limited comparability to real-world memory clinic environments.

The present study aimed to address this gap by thoroughly characterizing the disease stage-specific properties of MTL and parietal subregional atrophy markers in a biomarker-characterized, memory clinic-based sample. We tested how these markers, whether recorded cross-sectionally or longitudinally, develop with disease progression, utilizing both clinical as well as data-driven, continuous disease staging. The latter was used to gain insights into AD progression especially during early disease stages, where phenotypical changes are subtle and single biomarkers may be less reliable.^28^ Finally, we investigated the dynamic coupling of MRI readouts and scores on various established neuropsychological inventories for different potential uses, including diagnosis, prognosis, and monitoring at different disease stages.

## Methods

### Participants

We included data from 363 participants enrolled in the DZNE Longitudinal Cognitive Impairment and Dementia Study (DELCODE), stratified by their baseline diagnostic group and Aβ status.^29^ This sample consisted of Aβ-negative healthy controls (HC Aβ–, *n* = 165) and diagnostic groups reflecting the AD continuum according to the 2018 NIA-AA research criteria, including Aβ-positive healthy controls (HC Aβ+, *n* = 30), as well as Aβ-positive memory clinic patients with subjective cognitive decline (SCD Aβ+, *n* = 78), mild cognitive impairment (MCI Aβ+, *n* = 51), and mild dementia of the Alzheimer type (DAT Aβ+, *n* = 39).^30^ The full inclusion criteria are detailed in the Supplementary Methods.

Participants underwent baseline and annual follow-up assessments including MRI scanning and neuropsychological testing. Given our particular interest in MTL subregions, study visits were only considered if these data were available (see Supplementary Figure 1 for an overview of follow-up availabilities). This resulted in an average of 3.34 (*SD* = 0.81) total assessments per subject with an average follow-up interval of 1.08 (*SD* = 0.26) years between study visits in the analyzed sample. All participants provided their written informed consent to participate in the study in accordance with the Declaration of Helsinki. The local institutional review boards at all participating study sites approved the study protocol. DELCODE was registered with the German Clinical Trials Registry (DRKS; DRKS00007966) prior to inclusion of the first participants.

**Figure 1.**
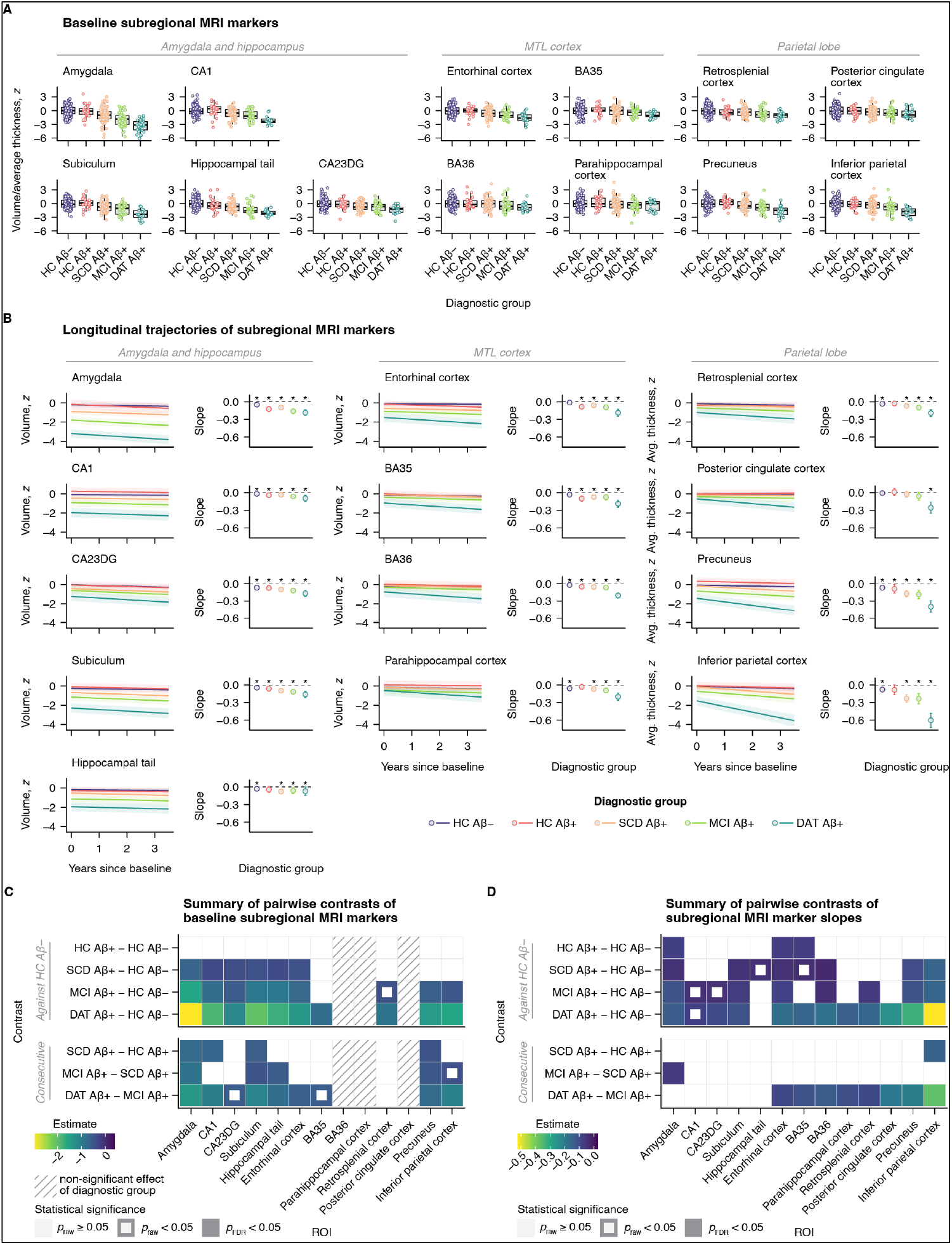
Baseline and longitudinal MTL and parietal subregional structure across the clinical AD continuum. (**A**) Boxplot diagrams of baseline gray matter volumes and average cortical thicknesses across regions of interest, grouped by meta-region. (**B**) Group-level longitudinal trajectories of structural markers (left panels) and the corresponding slopes (right panels), estimated from linear mixed-effects models. Ribbons and error bars denote 95% confidence intervals. Asterisks highlight significant slopes, i.e., significant change over time. (**C**) Matrix displaying estimates from ANCOVA *post hoc* group comparisons of baseline MRI markers. The striped pattern indicates measures with non-significant main effects of diagnostic group (not passed to *post hoc* tests). (**D**) Estimates from pairwise group comparisons of longitudinal slopes of subregional MRI markers. Abbreviations: BA, Brodmann area. CA, cornu ammonis. CA23DG, cornu ammonis 2, 3, and dentate gyrus. DAT, dementia of the Alzheimer type. FDR, false discovery rate. HC, healthy controls. IPC, inferior parietal cortex. MCI, mild cognitive impairment. MTL, medial temporal lobe. ROI, region of interest. SCD, subjective cognitive decline. SUB, subiculum.

### Fluid biomarkers

Data on Aβ-positivity were preferentially obtained from lumbar cerebrospinal fluid (CSF, *n* = 224, 61.71%). A threshold for Aβ-positivity of ≤0.08 was calculated through two-component Gaussian mixture modelling of Aβ_42_/Aβ_40_ ratios (Mesoscale Diagnostics LLC, Rockville, USA). If CSF was unavailable, we calculated individual probabilities of Aβ-positivity, ranging from 0 (least likely to be Aβ-positive in CSF) to 1 (most likely to be Aβ-positive in CSF). These probabilities were obtained using a binomial logistic regression model, predicting Aβ-positivity in CSF with age, APOE genotype, plasma phosphorylated-tau 181 concentration (Simoa assays; Quanterix, Billerica, MA, USA), and plasma Aβ_42_/Aβ_40_ ratios (Lumipulse G System assays; Fujirebio Inc., Tokyo, Japan; ref.^31^) included as predictors. A cutoff for positivity was identified using the Youden index and set at >0.639, with the cost ratio of false negative to false positive classifications set to 1.5:1. This decision was guided by evidence suggesting Aβ_42_/Aβ_40_ ratios to become abnormal earlier in CSF than in the peripheral blood, making specificity a higher priority than sensitivity.^32^ The model is further described in the Supplementary Methods and closely follows work by Hu *et al*.^33^

### Neuropsychological testing

We included a broad range of measures from the DELCODE neuropsychological testing battery, targeting different cognitive domains. This included tests of episodic memory (Free and Cued Selective Reminding Test–Free + Total Recall [FCSRT-96, ref.^34^], Wechsler Memory Scale IV [WMS-IV, ref.^35^] Logical Memory Delayed Recall, Alzheimer’s Disease Assessment Scale-Cognitive subscale [ADAS-Cog, ref.^36^] delayed and immediate word recall, Face Name Associative Recognition Task), language (sum of animals and groceries verbal fluency, FCSRT Naming), executive functions (Symbol-Digit Modalities Test [SDMT, ref.^37^], time difference of the Trail Making Test version B and A [TMT B–A], a Flanker task [ref.^38^], ADAS-Cog Number Cancellation), working memory (WMS-R Digit Span), and visuospatial functions (clock drawing and copying, ADAS-Cog Figure Savings and Copying). In addition, we assessed measures of clinical functioning (Clinical Dementia Rating Sum of Boxes scale [CDR-SB, ref.^39^], Functional Activities Questionnaire [FAQ, ref.^40^]), neuropsychiatric symptoms (Neuropsychiatric Inventory–Questionnaire [NPI-Q, ref.^41^]) and cognitive composite scores (ADAS-Cog-13, Mini-Mental State Examination [MMSE, ref.^42^], Preclinical Alzheimer Cognitive Composite [PACC-5, ref.^43^]). If visual inspection of the data indicated that test score distributions strongly deviated from normality, we applied pre-normalization using latent process modelling implemented in the “lcmm” R package.^44^ Cognitive test scores were *z*-standardized to participants without clinically significant cognitive impairment, i.e., the HC Aβ–, HC Aβ+, and SCD Aβ+ groups, and, if needed, inverted so that lower scores represented worse performance across inventories.

### MRI acquisition and segmentation procedure

MRI data were collected on 3 Tesla Siemens MRI systems, including a T1-weighted, 3D whole-brain magnetization prepared rapid gradient echo sequence (MPRAGE; TE/TR = 437/2500ms, inversion time = 1100ms, 7º flip angle, 1mm isotropic resolution) and a T2-weighted turbo spin-echo (TSE; TE/TR = 354/3500ms, 120° flip angle, 0.5 × 0.5 × 1.5mm resolution) sequences. To optimally visualize MTL subregions, TSE images were oriented orthogonally to the longitudinal axis of the hippocampus and were limited to a slab covering the MTL.^45^

MTL subregional volumes were derived using a newly developed longitudinal implementation of the automated segmentation of hippocampal subfields (ASHS, refs.^11,17^) algorithm, utilizing subject-specific templates. This pipeline can be applied to both T1-(T1-ASHS) and T2-weighted images (T2-ASHS). This flexibility is crucial since different structures are best segmented on different image modalities—for example, hippocampal subfields on T2-weighted and the amygdala on T1-weighted images.^46,47^ Technical details of this pipeline are summarized in the Supplementary Methods and were described in a previous publication from our group.^2^ T2-ASHS was used with the Penn ABC 3T atlas, delineating hippocampal subfields (cornu ammonis [CA] 1–3, dentate gyrus, subiculum) as well as the extrahippocampal MTL cortices (entorhinal cortex, Brodmann area [BA] 35 [approximately corresponding to Braak’s transentorhinal cortex, ref.^17^], BA36, and parahippocampal cortex).^19,48^ To reduce the number of regions of interest (ROIs), we combined the CA2, CA3, and dentate gyrus labels into a single CA23DG label, as these regions are less affected by early AD-related NFTs.^49^ Hippocampal subfields were only segmented in the anterior hippocampus (combining hippocampal head and body), while its posterior-most portion was segmented as hippocampal tail. T1-ASHS with an updated version of the PMC-T1 atlas was used to generate segmentations of the amygdala.^20,46^ We also conducted standard cross-sectional T2-ASHS segmentation of baseline images to evaluate the comparability of longitudinally derived volumes with those generated using the established T2-ASHS algorithm.

Average cortical thicknesses of the granular retrosplenial cortex (isthmus of the cingulate gyrus), inferior parietal cortex, posterior cingulate cortex, and the precuneus were generated from T1-weighted images using FreeSurfer’s longitudinal stream (v7.1.1, Desikan-Killiany atlas, http://surfer.nmr.mgh.harvard.edu/).^18^

Gray matter volumes were adjusted for their relationship with total intracranial volume in all HC Aβ– participants included in DELCODE.^50^ All structural measures were *z*-standardized to the HC Aβ– group. Pipeline availabilities are indicated in Supplementary Figure 2.

**Figure 2.**
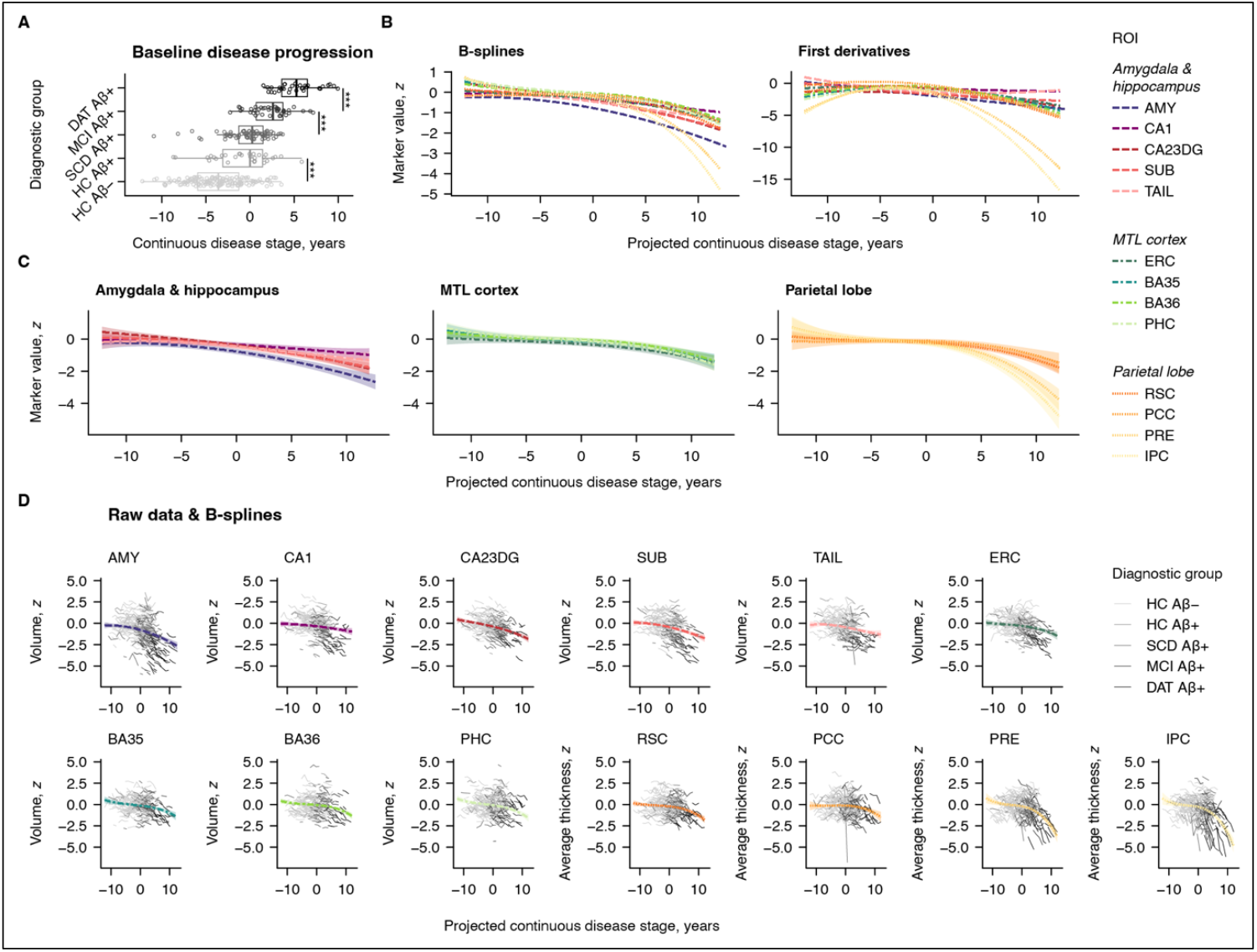
Non-linear changes in MTL and parietal subregional structure along a continuous estimate of disease progression, scaled in years. (**A**) Boxplot diagrams of baseline disease stage by diagnostic group, centered to the median of the HC Aβ+ group. (**B**) Cubic B-splines and their first derivatives were used to assess the development of structural MRI measures with disease progression. The estimated trajectories also shown in (**C**) with bootstrapped confidence intervals and clustered by meta-region. (**D**) For each ROI, the predicted curves also shown in (**B**) and (**C**) are plotted along with the observed participant-level trajectories. Abbreviations: Aβ, amyloid-β. AMY, amygdala. BA, Brodmann area. CA, cornu ammonis. CA23DG, cornu ammonis 2, 3, and dentate gyrus. DAT, dementia of the Alzheimer type. ERC, entorhinal cortex. HC, healthy controls. IPC, inferior parietal cortex. MCI, mild cognitive impairment. MTL, medial temporal lobe. PCC, posterior cingulate cortex. PHC, parahippocampal cortex. PRE, precuneus. ROI, region of interest. RSC, retrosplenial cortex. SCD, subjective cognitive decline. SUB, subiculum. TAIL, hippocampal tail.

### Data-driven continuous disease staging

A data-driven disease progression model was trained to capture disease progression on a continuous scale, closely following a previous analysis of DELCODE data.^51,52^ Briefly, the model was trained on all available longitudinal CSF Aβ_42_/Aβ_40_ ratios and phosphorylated tau 181 concentrations, as well as PACC-5 and ADAS-Cog-13 sum scores. While the previously reported model included entorhinal and hippocampal volume, we re-trained an otherwise analogous model in the same sample, this time excluding atrophy markers to avoid circularity in our analyses. Using this model, we estimated a continuous disease stage for each participant’s baseline study visit in reference to the entire DELCODE sample. To simplify interpretation, we corrected continuous disease stages for age, sex, and years of education and introduced a clinical reference through linear transformation, setting the median of the HC Aβ+ group to zero.

### Statistical analyses

Statistical analyses were performed in R v4.2.2.^53^ Two-way intraclass correlations tested agreement between baseline volumes from longitudinal and cross-sectional T2-ASHS. We employed ANCOVA to test for effects of study site on scaled MRI readouts within each diagnostic group. ANCOVAs were also used to compare the baseline MRI readouts across diagnostic groups. *Post hoc* comparisons comprised contrasts of all diagnostic groups against the HC Aβ– group and contrasts of diagnostic groups representing consecutive clinical disease stages (e.g., MCI Aβ+ *versus* SCD Aβ+). We investigated whether longitudinal structural trajectories differed across diagnostic groups through linear mixed effects models with participant-level random slopes and intercepts and applied the same contrasts as described above.

Next, ANOVA was used to test for group differences in disease stage at baseline. To model atrophy trajectories along continuous disease stages, we estimated longitudinal disease stages by adding years since baseline to each individual’s baseline estimate of disease stage. These projected stages were used in linear mixed-effects models to predict each atrophy marker. Cubic B-splines (*df* = 3) were used to capture non-linear mean trajectories, and the first derivatives of the resulting curves were examined to assess slope changes with disease progression.

Finally, we analyzed the longitudinal relationships of regional structure and cognitive measures using bivariate latent growth curve models (LGCMs) implemented in the “OpenMx” R package.^54,55^ Bivariate LGCMs allow for the estimation of change in two longitudinally recorded variables and the relationships thereof. Models were specified and fitted for each pairing of structural and cognitive variables as visualized in Supplementary Figure 3. An acceptable model fit was defined by a comparative fit index (CFI) > 0.90 and a root mean square error of approximation (RMSEA) < 0.08. Wald tests (critical *Z* = 1.96) were used to ensure sufficient variance in latent intercepts and slopes to test for covariance. Likelihood-ratio tests were used to freely estimate covariance parameters of interest, each reflecting different potential use cases for structural MRI markers. This included the baseline-baseline (diagnosis and case finding), baseline-slope (prognosis) and slope-slope (disease monitoring) pairwise associations of structural and cognitive variables. We also investigated if these relationships differed across clinical disease stages (HC Aβ–, preclinical AD comprising CU+ and SCD Aβ+, and symptomatic AD comprising MCI Aβ+ and DAT Aβ+). To this end, multi-group models were estimated where the parameter of interest was freely estimated within each group, constraining all other parameters to be equal across groups. Group-wise parameters were tested against 0 within each group using likelihood-ratio tests.

**Figure 3.**
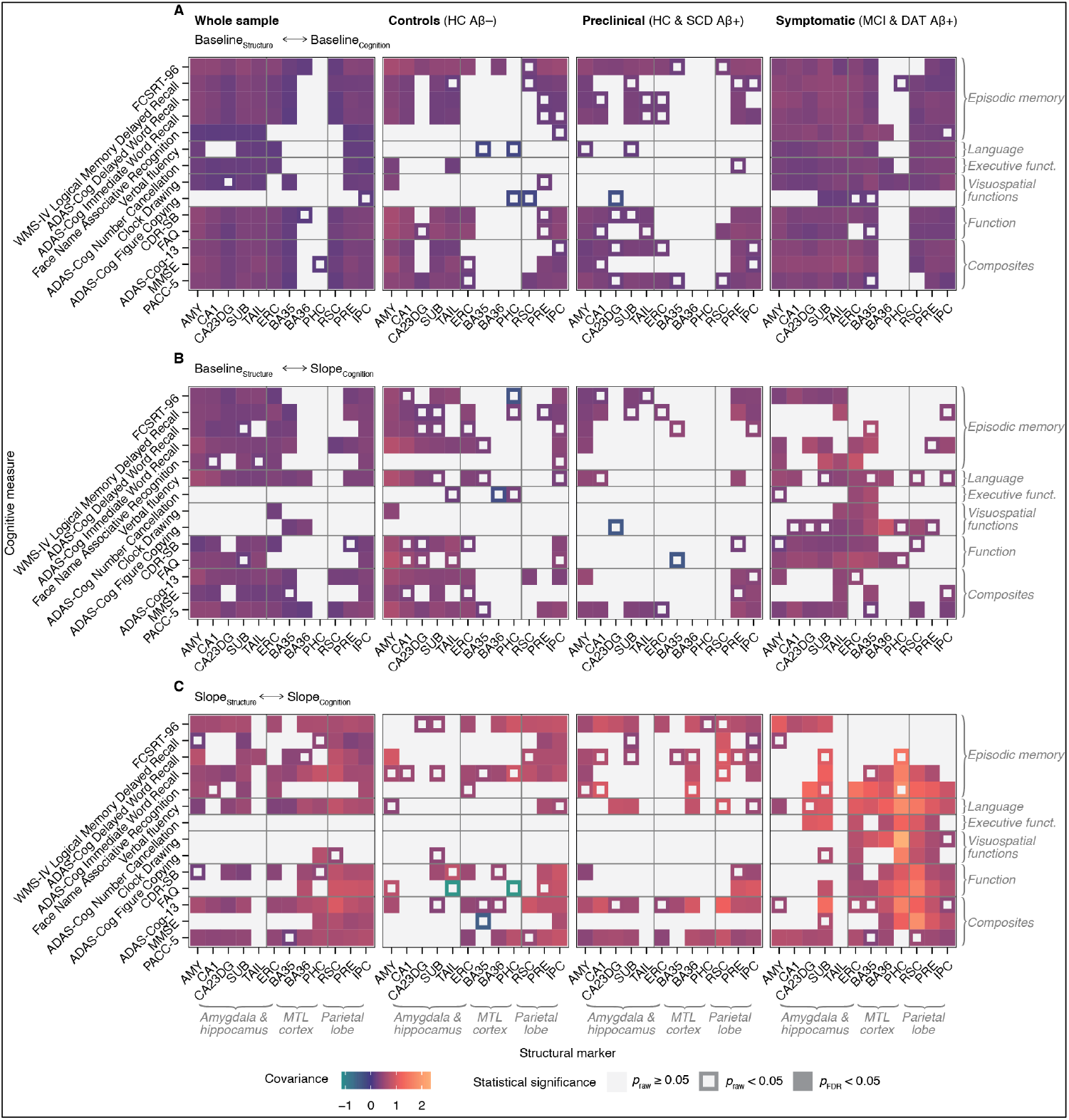
Estimates from bivariate LGCMs capturing the disease stage-specific cross-sectional and longitudinal relationships of subregional structural MRI measures and cognitive test scores. The covariance parameters of interest were the (**A**) baseline-baseline (**B**) baseline-slope, and (**C**) slope-slope pairwise associations of ROI structure and cognitive score. Abbreviations: AMY, amygdala. ADAS-Cog, Alzheimer’s Disease Assessment Scale-Cognitive subscale. BA, Brodmann area. CA, cornu ammonis. CA23DG, cornu ammonis 2, 3, and dentate gyrus. CSD-SB, Clinical Dementia Rating Sum of Boxes scale. ERC, entorhinal cortex. FAQ, Functional Activities Questionnaire. FCSRT, Free and Cued Selective Reminding Test. FDR, false discovery rate. IPC, inferior parietal cortex. MMSE, Mini-Mental State Examination. MTL, medial temporal lobe. PACC-5, Preclinical Alzheimer Cognitive Composite. PHC, parahippocampal cortex. PRE, precuneus. RSC, retrosplenial cortex. SUB, subiculum. TAIL, hippocampal tail.

All analyses controlled for baseline age and sex. Years of education was included as a covariate when predicting cognitive scores. Models including disease time were not corrected for demographics as they were regressed out of this metric. The threshold for statistical significance was *p* ≤.05 and false discovery rate (FDR) correction was applied where appropriate.

## Results

An overview of participant characteristics is provided in Table 1. Intra-class correlation coefficients of baseline volumes generated using the standard cross-sectional and in-house longitudinal T2-ASHS pipelines were excellent, supporting their comparability (*n* = 272, mean ICC = 0.94, *SD* = 0.02; range = 0.90 – 0.97, all *p* <.001; Supplementary Figure 4). There were no statistically significant effects of site on scaled structural markers across ROIs and diagnostic group (Supplementary Table 1, Supplementary Figure 5).

**Table 1.** Characteristics of the analyzed sample.

|  | Missing<br>*n* (%) | Diagnostic group |  |  |  |  | Total<br>*N* = 363 |
| --- | --- | --- | --- | --- | --- | --- | --- |
| | | HC Aβ-<br>*n* = 165 | HC Aβ+<br>*n* = 30 | SCD Aβ+<br>*n* = 78 | MCI Aβ+<br>*n* = 51 | DAT Aβ+<br>*n* = 39 | |
| Baseline age, years | – | 68.14 (5.05) | 71.23<br>(6.51) | 73.07<br>(5.55) | 73.75<br>(4.82) | 74.74<br>(6.58) | 70.95 (6.04) |
| Sex, female | – | 106<br>(64.24%) | 12<br>(40.00%) | 29<br>(37.18%) | 27<br>(52.94%) | 19<br>(48.72%) | 193<br>(53.17%) |
| Education level,<br>years | – | 14.55 (2.78) | 14.27<br>(3.14) | 14.50<br>(2.98) | 14.08<br>(3.32) | 12.97<br>(2.86) | 14.28 (2.96) |
| Baseline disease<br>stage, years | 33<br>(9.09%) | –3.61 (3.26) | –0.92<br>(3.35) | –0.27<br>(2.63) | 2.32 (2.16) | 5.34 (2.31) | –0.93 (4.17) |
| Number of<br>assessments | *n.a.* | 3.56 (0.70) | 3.67 (0.66) | 3.17 (0.83) | 3.18 (0.87) | 2.72 (0.76) | 3.34 (0.81) |
| **Pipeline availability** |  |  |  |  |  |  |  |
| T1-ASHS | *n.a.* | 148<br>(89.70%) | 27<br>(90.00%) | 76<br>(97.44%) | 50<br>(98.04%) | 37<br>(94.87%) | 338<br>(93.11%) |
| T2-ASHS | *n.a.* | 151<br>(91.52%) | 28<br>(93.33%) | 61<br>(78.21%) | 32<br>(62.75%) | 18<br>(46.15%) | 290<br>(79.89%) |
| FreeSurfer | *n.a.* | 165<br>(100.00%) | 30<br>(100.00%) | 78<br>(100.00%) | 51<br>(100.00%) | 39<br>(100.00%) | 363<br>(100.00%) |
| **Cognitive test scores\*** |  |  |  |  |  |  |  |
| PACC-5 | 37<br>(10.19%) | 0.23 (0.54) | –0.16<br>(0.74) | –0.36<br>(0.71) | –1.28<br>(0.72) | –3.52<br>(1.35) | –0.30 (1.11) |
| MMSE | 1 (0.28%) | 29.57 (0.71) | 29.20<br>(1.06) | 29.17<br>(1.04) | 27.84<br>(1.60) | 23.51<br>(3.36) | 28.56 (2.34) |
| ADAS-Cog-13 | 9 (2.48%) | 5.34 (3.01) | 6.92 (4.22) | 8.19 (3.93) | 16.50<br>(6.63) | 30.22<br>(7.66) | 10.08 (8.86) |
| CDR-SB | 7 (1.93%) | 0.05 (0.21) | 0.26 (0.85) | 0.51 (0.76) | 1.36 (0.96) | 4.29 (1.95) | 0.80 (1.54) |

\*Raw scores are displayed. Abbreviations: ADAS-Cog-13, Alzheimer's Disease Assessment Scale-Cognitive subscale sum score. ASHS, automated segmentation of hippocampal subfields. CDR-SB, Clinical Dementia Rating Sum of Boxes. DAT, dementia of the Alzheimer type. HC, healthy control. MCI, mild cognitive impairment. MMSE, Mini-Mental State Examination. *n.a.*, not applicable. PACC-5, Preclinical Alzheimer Cognitive Composite. SCD, subjective cognitive decline.

### Baseline assessments reveal atrophy of early Braak regions in preclinical Alzheimer’s disease with subjective cognitive decline

The first main analysis aimed to identify potential baseline differences of atrophy markers across diagnostic groups using ANCOVAs (Figure 1A, C, see also Supplementary Tables 2 and 3). Structural MRI readouts from Aβ-positive groups were first compared to those of HC Aβ– participants. We found no significant differences for the HC Aβ+ group. Meanwhile, volumes of the amygdala (*b* = –0.70, *t* = –3.75, *p*_FDR_ < .001), the entorhinal cortex (*b* = –0.44, *t* = –2.75, *p*_FDR_ = .015), and all hippocampal ROIs (all *b* ≤ –0.36, *t* ≤ –2.14, *p*_FDR_ ≤ .047) were significantly reduced in the SCD Aβ+ group. The same regional volumes were reduced in MCI Aβ+ participants, though to a numerically greater extent (all *b* ≤ –0.59, *t* ≤ –2.97, *p*_FDR_ ≤ .011). In addition, MCI Aβ+ participants exhibited significantly reduced cortical thickness in the precuneus (*b* = –0.62, *t* = –2.53, *p*_FDR_ = .028) and in the inferior parietal cortex (*b* = –0.51, *t* = –2.45, *p*_FDR_ = .035). All structural markers included in the *post hoc* analysis were significantly reduced in DAT Aβ+ patients (all *b* ≤ –0.80, *t* ≤ –3.18, *p*_FDR_ ≤ .011). Analysing further contrasts representing consecutive diagnostic stages, we observed significant differences across comparisons (SCD Aβ+ *versus* HC Aβ+, MCI Aβ+ *versus* SCD Aβ+, DAT Aβ+ *versus* MCI Aβ+). These were mainly focused on the amygdala, hippocampal ROIs, the precuneus and the inferior parietal cortex.

Diagnostic group had a non-significant effect on BA36 and parahippocampal cortex volume, as well as on posterior cingulate cortex thickness. These models were not passed to *post hoc* analysis.

### Longitudinal MRI reveals accelerated atrophy in preclinical Alzheimer’s disease

Now also considering follow-up data, we tested if diagnostic groups differed in their longitudinal trajectories of structural readouts using linear mixed effect models. Random slopes showed insufficient variance for when predicting posterior cingulate cortical thickness and were therefore omitted. Group-wise trajectories over time as well as their associated slope (i.e., annual rate of change) estimates are displayed in Figure 1B. Figure 1D provides a summary of pairwise group comparisons of slopes (see also Supplementary Tables 4–6). In HC Aβ– participants, a significant decline was observed across markers except for entorhinal volume and posterior cingulate cortical thickness.

Decline was further amplified in several regions within the different Aβ-positive groups. While no significant effects were observed for the HC Aβ+ group in the cross-sectional analysis, there was an accelerated volumetric decline over time in the amygdala (*b* = –0.07, *t* = –2.99, *η*^2^_partial_ = 0.03, *p*_FDR_ = .007), entorhinal cortex (*b* = –0.07, *t* = –3.01, *η*^2^_partial_ = 0.04, *p*_FDR_ = .007), and BA35 (*b* = –0.07, *t* = –2.73, *η*^2^_partial_ = 0.03, *p*_FDR_ = .016). The SCD Aβ+ group exhibited an even more pronounced structural decline relative to the HC Aβ– group, particularly in parietal subregions (precuneus: *b* = –0.10, *t* = –3.10, *η*^2^_partial_ = 0.04, *p*_FDR_ = .005; inferior parietal cortex: *b* = –0.15, *t* = –3.89, *η*^2^_partial_ = 0.06, *p*_FDR_ < .001). Additional regions within the medial temporal lobe (amygdala, subiculum, entorhinal cortex, and BA36) also showed significantly faster decline in the SCD Aβ+ group. When comparing SCD Aβ+ and HC Aβ+ participants, the only significant difference was in the rate of thinning in the inferior parietal cortex (*b* = –0.15, *t* = –2.74, *η*^2^_partial_ = 0.03, *p*_FDR_ = .009). A similar pattern to that observed in SCD Aβ+ pattern to that observed in SCD Aβ+ individuals was found in the MCI Aβ+ group, with an additional significant acceleration of thinning over time in the retrosplenial cortex (*b* = –0.06, *t* = –2.67, *η*^2^_partial_ = 0.03, *p*_FDR_ = .027). In DAT Aβ+ participants, nearly all structures, except for the hippocampal tail and CA1, showed accelerated atrophy. Compared to MCI Aβ+ participants, structural decline in DAT Aβ+ individuals progressed more rapidly in extrahippocampal and parietal regions, while hippocampal and amygdalar structures showed no significant differences in atrophy rates.

### Data-driven disease staging offers detailed insights into subregional atrophy progression

Stepping away from discrete classifications, we next analyzed the evolution of regional atrophy readouts in relation to continuous, data-driven disease stages. The disease progression model used to derive participant-level disease stages is illustrated in Supplementary Figure 6. We found that baseline disease stage increased with clinical progression (i.e., with each successive level of diagnostic group), except that there was no significant difference between HC Aβ+ and SCD Aβ+ participants (*b* = 0.65, *p*_FDR_ = 0.308; otherwise range (*b*): 2.58 – 3.02, all *p*_FDR_ < .001; Figure 2A). Linear mixed effects models with B-splines were used to capture non-linearity in the longitudinal trajectories of structural MRI readouts with continuous disease stage. Figures 2 B–D show the fitted slopes and their first derivatives. As expected in a sample of older adults, progressive atrophy (negative first derivatives) was evident already at early stages below the HC Aβ+ group median, which was set to be zero. Within the MTL, the amygdala exhibited the most pronounced decline with disease progression, followed by the MTL cortex were relatively small, with a modest acceleration occurring later, around the median disease stage in DAT Aβ+ participants. A similar pattern was observed for parietal gray matter thickness in the early disease stages. However, as the disease advanced to stages predominantly seen in symptomatic individuals, a rapid acceleration became evident, particularly in the precuneus and inferior parietal cortex. Interestingly, the first derivatives of the fitted curves indicate that hippocampal and amygdalar atrophy markers stabilize in the late disease stages. In contrast, the decline of atrophy markers in the MTL cortex— and even more so in the parietal lobe—continued to accelerate as modeled disease stages approached their maximum.

### Structural and cognitive changes covary uniquely in different disease stages

Finally, we analyzed longitudinal measures of regional brain structure and cognition in order to identify their potentially disease stage-specific couplings. To this end, bivariate LGCMs were fitted both in the whole sample and in sub-samples representing different clinical disease stages. Due to their non-normal distributions, ADAS-Cog Delayed Word Recall, ADAS-Cog Figure Copying, ADAS-Cog-13, CDR-SB, Clock Copying, Clock Drawing, FAQ, MMSE, and NPI-Q scores were pre-normalized using latent process modelling (Supplementary Results). Most models showed adequate fit and variance structures. However, FCSRT Naming SDMT, TMT B–A, Flanker, WMS-R Digit Span, Clock Copying, and NPI-Q scores, as well as posterior cingulate cortical thicknesses had insufficient slope variances and are therefore not reported. Additional details on model fit are reported in the Supplementary Results and Supplementary Figures 7 and 8. The estimates for all covariance parameters of interest are visualized in Figure 3.

As expected, all significant covariance estimates were positive, indicating that structural decline was linked to worsening cognition. However, no consistent covariance pattern emerged across diagnostic groups; instead, each group showed distinct associations between subregional atrophy markers and individual test scores.

Group-specific covariance matrices revealed that significant estimates were more frequent and descriptively higher in the symptomatic group (MCI Aβ+ and DAT Aβ+) compared to the HC Aβ– and preclinical (HC Aβ+ and SCD Aβ+) groups, reflecting stronger structure-cognition coupling at later clinical stages. Structure-cognition relationships were numerically weaker in cross-sectional analyses (baseline-baseline, Figure 3A) than in analyses where structure was assessed cross-sectionally and cognition longitudinally (baseline-slope, Figure 3B). Covariances were numerically largest when both structure and cognition were assessed longitudinally (slope-slope, Figure 3C), suggesting that brain structure and cognition are more closely linked through their changes over time than through their baseline measures. A notable shift was observed in regional patterns when comparing baseline-slope and slope-slope associations: when brain structure is considered longitudinally instead of cross-sectionally, significant covariances are increasingly focused on the structures in the MTL cortices and the parietal lobe. This pattern occurred across groups but was most pronounced in symptomatic participants.

In preclinical stages, the FCSRT-96 and PACC-5 demonstrated the most consistent associations across covariance parameters and regions, with the FCSRT-96 showing more widespread and significant structural correlates than other episodic memory measures. Significant slope-slope associations were also observed in models including the ADAS-Cog-13. Functional decline was linked to thinning in the precuneus and inferior parietal cortex, while associations with language, executive functioning, or visuospatial scores were rare.

In symptomatic groups, cross-sectional associations between structural and cognitive measures were notably broad, with only parahippocampal cortex and BA36 volumes showing limited associations. Baseline-slope relationships were primarily centered on MTL structures across cognitive domains, while slope-slope covariances focused on the MTL cortey and parietal lobe, encompassing cognitive domains beyond episodic memory and cognitive composite.

## Discussion

This study provides a detailed characterization of MTL and parietal subregional atrophy markers across the AD spectrum. Using specialized longitudinal segmentation techniques, we identified accelerated atrophy rates in MTL subregions already in preclinical AD. SCD in cognitively unimpared individuals was associated with widespread, manifest atrophy. Meanwhile, symptomatic disease stages were characterized especially by increasing atrophy of the hippocampus and parietal cortices. Independent of clinical diagnoses, the examined atrophy markers uniquely developed within a data-driven disease staging framework, with the amygdala showing earliest decline. Addressing different contexts-of-use, we provide insights into associations of atrophy markers and cognition by analyzing a comprehensive set of cross-sectionally and longitudinally recorded neuropsychological inventories.

### Accelerated atrophy rates are detectable in preclinical Alzheimer’s disease

The atrophy patterns identified across both clinical and data-driven disease stages mostly mirror the established Braak staging framework of NFT progression, with the MTL being affected prior to the parietal cortex.^8,12^ This aligns with previous findings on the co-localization of NFTs and atrophy, as well as studies on MRI signatures of AD, further supporting the use of structural MRI as a biomarker for AD-related neurodegeneration.^3,7–9,56^

Highlighting significant variations in the onset and progression of atrophy across small anatomical structures, our study underscores the importance of subregional analyses, particularly when seeking to maximize the MRI sensitivity during early disease stages. For instance, our longitudinal data suggest that in preclinical AD, atrophy spares the posterior MTL (parahippocampal cortex, hippocampal tail), while anterior structures deteriorate earlier.

Going into more anatomical detail within the MTL, accelerated rates of atrophy in hippocampal subfields emerged only after the anterior MTL cortices (entorhinal cortex, BA35, BA36) and the amygdala were affected. These structures showed an abnormally fast volumetric decline as early as AD stage 1 per NIA-AA 2018 criteria (HC Aβ+).^30^ While the MTL cortices are known as early hotspots of AD-related NFTs and atrophy, the amygdala has been underrepresented in relevant research.^57^ Our study showed declining amygdalar volumes already during earliest disease stages, suggesting that it may be more sensitive to early AD than the commonly used hippocampal volume. Complementing these findings, previous studies have reported longitudinal atrophy in the amygdala, transentorhinal (approximately corresponding to BA35), and entorhinal regions in cognitively unimpaired older adults who later converted to MCI and earlier abnormal atrophy in the entorhinal cortex and amygdala than in the hippocampus.^58,59^ This effect likely reflects early AD-related NFTs but may also result from non-AD proteinopathies (e.g., TDP-43 or α-synuclein) accumulating in the amygdala.^57^ Interestingly, hippocampal and amygdalar atrophy rates stabilized towards later data-driven disease stages and did not differ between DAT Aβ+ and MCI Aβ+ participants, indicating reduced sensitivity in symptomatic AD.

### Subjective cognitive decline is associated with manifest atrophy in preclinical Alzheimer’s disease

Our cross-sectional data provides evidence of widespread, manifest neurodegeneration in the MTL among SCD Aβ+ but not HC Aβ+ individuals (i.e., in stage 2 but not stage 1 AD per NIA-AA criteria). In addition, our longitudinal analyses revealed abnormally fast atrophy rates in the precuneus and inferior parietal cortex in SCD Aβ+ but not HC Aβ+ participants. Although some studies have linked SCD to parietal atrophy, there is no clear consensus on its atrophy correlates, likely due to varied methodologies, diverging segmentation procedures, and inconsistent SCD definitions.^60^ The more extensive atrophy in SCD Aβ+ compared to HC Aβ+ participants aligns with studies linking SCD in preclinical AD to elevated biomarkers of AD proteinopathies and an elevated risk of progression to cognitive impairment.^29,61–64^

### Cross-sectional and longitudinal atrophy markers provide distinct insights into disease progression

Our study shows that cross-sectionally and longitudinally recorded MRI markers provide distinct, yet complementary, insights into accumulated and ongoing regional atrophy, respectively. For example, in SCD Aβ+ participants, precuneus and inferior parietal cortical thickness appeared normal cross-sectionally but showed accelerated atrophy over time in longitudinal assessments. Meanwhile, significant accumulated atrophy of these regions became evident in the subsequent clinical disease stage (MCI Aβ+). While most findings align with this pattern of accelerated decline (ongoing atrophy) preceding cross-sectional reductions (accumulated atrophy), some results deviate from this trend. For example, our cross-sectional analyses identified reduced volumes in hippocampal subfield CA1 at the SCD Aβ+ stage while none of the diagnostic groups showed abnormally accelerated CA1 atrophy rates over time. Importantly, we believe it is not paradoxical that atrophy rates in BA35, BA36, and the parahippocampal cortex accelerate during preclinical stages despite the absence of significant cross-sectional abnormalities before the DAT Aβ+ stage. This can be attributed to the high inter-individual macroanatomical differences in these structures, which adds variance to absolute volumetric measures unrelated to neurodegeneration.^17^ Longitudinal within-subject volumetric analyses account for this variance, making them better suited to capture subtle atrophy in these structures.^18^

### Brain structure and cognition show disease stage-specific couplings

The covariance patterns between MRI markers and neuropsychological scores consistently linked atrophy to cognitive decline, but the measures involved and their strength of association varied across parameters. These parameters represented distinct uses for biomarkers: baseline-baseline covariances are relevant for diagnostic and case finding purposes, baseline-slope relationships reflect the prognostic value of an atrophy marker, and slope-slope associations indicate an atrophy marker’s suitability for monitoring disease progression. Hence, our data suggest that the intended use, analytical design, and neuropsychological correlates of interest should be considered when selecting an MRI readout. Overall, however, our findings suggest that atrophy-cognition associations are strongest when assessed longitudinally rather than cross-sectionally, aligning with previous studies.^65–67^

While focusing on AD, our study also provides insights into brain structure-cognition relationships in healthy aging (HC Aβ–). Contrasting earlier studies, we found no widespread longitudinal relationships between MTL structure and cognition, particularly episodic memory.^68^ This may reflect a lag between atrophy and cognitive decline, as indicated by broader associations of baseline MTL structure with future cognitive decline.

Building on previous reports of varying sensitivities of cognitive inventories with AD progression, our results highlight that the associations of regional brain atrophy and neuropsychological test scores differ in preclinical and symptomatic AD.^69,70^ For instance, entorhinal volume, a commonly used atrophy marker in AD, was longitudinally related to clinical functioning in symptomatic but not preclinical disease stages. Regarding associations of specific atrophy markers and test scores, our results largely overlap with existing neuropsychological and -pathological literature by both capturing established anatomoclinical relationships and highlighting their dynamic fluctuations over the disease course. For example, cognitive decline was more strongly linked to emerging parietal atrophy in symptomatic than in preclinical AD, aligning with reports of increasing NFT pathology in the parietal lobe as the disease progresses.^8,71^ In addition, the significant slope-slope relationships of FCSRT-96 and verbal fluency scores with MTL volumes in preclinical AD complement the established role of the MTL in episodic and semantic memory.^56,72,73^ It was especially the FCSRT-96 that stood out with consistent associations with MTL structure. This is unsurprising, as previous studies highlighted its strong association with early tau burden in preclinical AD as well as its relative importance among PACC-5 subtests for distinguishing preclinical AD from healthy aging over time.^43,74^ In contrast, many episodic memory measures, such as the ADAS-Cog Delayed Word Recall and WMS-IV Logical Memory Delayed Recall, showed weak associations with MTL structure. Among cognitive composites, the MMSE was largely unrelated to atrophy in preclinical AD, while significant associations were found for the ADAS-Cog-13 and PACC-5. These composites were designed to specifically capture AD-related cognitive decline and emphasize episodic memory, which may explain their relatively early associations with MTL atrophy.

### Limitations

Our study should be interpreted given its limitations. First, we did not account for potential atrophy heterogeneity, which has been reported previously.^2,75^ Second, although the atrophy patterns identified in the current study align with Braak staging, we were unable to test if NFTs, or other pathologies, indeed underlie local atrophy—distant effects are also possible.^7^ Third, our analyses of structure-cognition relationships cannot answer questions of possible mediating or moderating mechanisms, including functional alterations and neuroinflammatory responses. Fourth, disparities in sample sizes across diagnostic groups may have led to underpowered analyses, emphasizing the importance of the reported effect sizes. Finally, future studies should aim to replicate our findings in independent, more diverse cohorts.

### Conclusion

Our study offers detailed insights into the dynamics of anatomically-detailed atrophy markers within the MTL and parietal lobe across the AD continuum. We show that abnormalities on structural MRI do not necessarily appear imminent to cognitive impairment onset. Instead, accelerated volumetric decline in the entorhinal cortex, BA35, and the amygdala was observed already in preclinical AD. However, these markers appear less effective during symptomatic stages, where disease progression leads to more pronounced atrophy in hippocampal and parietal subregions. We demonstrate that atrophy-cognition associations are disease stage-specific and vary by context-of-use. Crucially, these associations were highly heterogeneous even within broader brain regions and cognitive domains, such as among MTL subregions and episodic memory tests. These results indicate that achieving optimal use of structural MRI in settings including clinical practice or intervention trials, requires readouts that are tailored to context-specific factors, including the intended use case and the targeted disease stage.

## Data Availability

The data, which support this study, are not publically available, but may be provided upon reasonable request.

## Acknowledgements

We thank Sarah Kriener and Merle Hinz for their support in preparing the MTL subregional segmentations, as well as Anke Jahn-Brodmann for her expert technical assistance.

