## Supplementary for "Disease stage-specific atrophy markers in Alzheimer’s disease"

### Supplementary material

#### DELCODE study group

##### Group members

| Name | Affiliation | Name | Affiliation | Name | Affiliation |
|---|---|---|---|---|---|
| Holger Amthauer | 21 | Ina Vogt | 4 | Daniela Frimmer | 16 |
| Arda Can Cetindag | 2 | Michael Wagner | 4,6 | Brigitte Huber | 16 |
| Nicoleta Carmen Cosma | 2 | Steffen wolfsgruber | 4 | Daniel Janowitz | 16 |
| Dominik Diesing | 2 | Claudia Bartels | 9 | Max Kreuzer | 16 |
| Marie Ehrlich | 2 | Peter Dechent | 27 | Claudia Müller | |
| Ersin Ersözlü | 1,2 | Niels Hansen | 9 | Axel Rominger | |
| Frederike Fenski | 2 | Lina Hassoun | 9 | Jennifer Schmid | 16 |
| Silka Dawn Freiesleben | 2 | Sina Hirschel | 9 | Anna Seegerer | 16 |
| Manuel Fuentes | 1,2 | Sabine Nuhn | | Julia Stephan | |
| Dietmar Hauser | 2 | Ilona Pfahlert | 9 | Adelgunde Zollver | 16 |
| Nicole Hujer | 2 | Lena Rausch | | Lena Burow | 23 |
| Enise Irem Incesoy | 2 | Björn Schott | 8,9,60 | Sylvia de Jonge | 23 |
| Christian Kainz | 22 | Jens Wiltfang | 8,9 | Peter Falkai | 23 |
| Catharina Lange | 2 | Heike Zech | 8,9 | Natalie Garcia Angarita | 23 |
| Katja Lindner | 2 | Abdelmajid Bader | 10 | Thomas Görlitz | 23 |
| Herlind Megges | 1,2 | Juan Carlos Baldermann | 10 | Selim Üstün Gürsel | 23 |
| Oliver Peters | 1,2 | Alexander Drzezga | 35 | Ildiko Horvath | 15 |
| Lukas Preis | 2 | Nasim Roshan Ghiasi | 10 | Carolin Kurz | 23 |
| Slawek Altenstein | 1,3 | Katja Hardenacke | 10 | Eva Meisenzahl-Lechner | |
| Andrea Lohse | | Frank Jessen | 4,1 | Robert Perneczky | 15,23,24,25 |
| Christiana Franke | 3 | Hannah Lützerath | 10 | Julia Utecht | 23 |
| Josef Priller | 1,3 | Franziska Maier | 10 | Martin Dyrba | 17 |
| Eike Spruth | 3 | Anja Martikke | 10 | Heike Janecek-Meyer | 18 |
| Miriam Barkhoff | 4 | Dix Meiberth | 10 | Ingo Kilimann | 17 |
| Henning Boecker | 4 | Ayda Rostamzadeh | 10 | Chris Lappe | 18 |
| Frederic Brosseron | 4 | Lena Sannemann | 10 | Esther Lau | 17 |
| Marcel Daamen | 4 | Ann-Katrin Schild | 10 | Henrike Pfaff | 18 |
| Tanja Engels | 4 | Susanne Sorgalla | 10 | Petr Sabik | 17 |
| Jennifer Faber | 4,7 | Simone Stockter | 10 | Monika Schmidt | 17 |
| Klaus Fließbach | 4 | Manuela Thelen | 10 | Heike Schulz | 17 |
| Ingo Frommann | 4 | Maike Tscheuschler | 10 | Sarah Schwarzenboeck | |
| Marcus Grobe-Einsler | 4,7 | Franziska Uhle | 10 | Stefan Teipel | 17,18 |
| Guido Hennes | 4 | Philip Zeyen | 10 | Marc-Andre Weber | |
| Gabi Herrmann | 4 | Laura Dobisch | 11 | Martina Buchmann | |
| Lorraine Jost | 4 | Emrah Düzel | 11 | Tanja Heger | 20 |
| Pascal Kalbhen | 6 | Doreen Grieger-Klose | 11 | Petra Hinderer | 19 |
| Okka Kimmich | 4,7 | Deike Hartmann | 11 | Elke Kuder-Buletta | 19 |
| Xenia Kobeleva | 4 | Coraline Metzger | 11 | Christoph Laske | 19,2 |
| Barbara Kofler | | Christin Ruß | 11 | Matthias Munk | 19,2 |
| Cornelia McCormick | 4 | Franziska Schulze | 11 | Christian Mychajliw | 19,2 |
| Lisa Miebach | 4 | Oliver Speck | 11 | Sebastian Sodenkamp | 19,50 |
| Carolin Miklitz | 4 | Glanz Wenzel | 11 | Surjo Soekadar | |
| Demet Oender | 4,7 | Renat Yakupov | 11 | Patricia Sulzer | 20 |
| Sandra Röske | 4 | Gabriel Ziegler | 11 | Theresia Trunk | |
| Christine Schneider | 4 | Katharina Bürger | 15,16 | Luisa Schneider | 1,2,58,61 |
| Anja Schneider | 4,6 | Lisa Coloma Andrews | 16 | Xiao Wang | 1,2,58,61 |
| Annika Spottke | 4,7 | Martin Dichgans | 15,16 | Kai Shao | 4,62 |
| Melina Stark | 4,6 | Birgit Ertl-Wagner | 26 | | |

#### Group affiliations


1 German Center for Neurodegenerative Diseases (DZNE), Berlin, Germany

2 Charité – Universitätsmedizin Berlin, corporate member of Freie Universität Berlin and Humboldt-Universität zu Berlin-Institute of Psychiatry and Psychotherapy

3 Department of Psychiatry and Psychotherapy, Charité, Charitéplatz 1, 10117 Berlin, Germany

4 German Center for Neurodegenerative Diseases (DZNE), Bonn, Venusberg-Campus 1, 53127 Bonn, Germany

5 Department of Psychiatry and Psychotherapy, University of Bonn, Venusberg-Campus 1, 53127 Bonn, Germany

6 Department of Old Age Psychiatry and Cognitive Disorders, University Hospital Bonn and University of Bonn, Bonn, Germany

7 Department of Neurology, University of Bonn, Venusberg-Campus 1, 53127 Bonn, Germany

8 German Center for Neurodegenerative Diseases (DZNE), Goettingen, Germany

9 Department of Psychiatry and Psychotherapy, University Medical Center Goettingen, University of Goettingen, Von-Siebold-Str. 5, 37075 Goettingen

10 Department of Psychiatry, University of Cologne, Medical Faculty, Kerpener Strasse 62, 50924 Cologne, Germany

11 German Center for Neurodegenerative Diseases (DZNE), Magdeburg, Germany

12 Institute of Cognitive Neurology and Dementia Research (IKND), Otto-von-Guericke University, Magdeburg, Germany

13 Department of Psychiatry and Psychotherapy, Otto-von-Guericke University, Magdeburg, Germany

14 Department of Neurology, Otto-von-Guericke University, Magdeburg, Germany

15 German Center for Neurodegenerative Diseases (DZNE, Munich), Feodor-Lynen-Strasse 17, 81377 Munich, Germany

16 Institute for Stroke and Dementia Research (ISD), University Hospital, LMU Munich, Feodor-Lynen-Strasse 17, 81377 Munich, Germany

17 German Center for Neurodegenerative Diseases (DZNE), Rostock, Germany

18 Department of Psychosomatic Medicine, Rostock University Medical Center, Gehlsheimer Str. 20, 18147 Rostock

19 German Center for Neurodegenerative Diseases (DZNE), Tübingen, Germany

20 Section for Dementia Research, Hertie Institute for Clinical Brain Research and Department of Psychiatry and Psychotherapy, University of Tübingen, Tübingen, Germany

21 Charité – Universitätsmedizin Berlin, corporate member of Freie Universität Berlin, Humboldt-Universität zu Berlin, and Berlin Institute of Health, Department of Nuclear Medicine, Augustenburger Platz 1, 13353 Berlin

22 Center for Cognitive Neuroscience Berlin (CCNB), Department of Education and Psychology, Freie Universität Berlin, Berlin, Germany

23 Department of Psychiatry and Psychotherapy, University Hospital, LMU Munich, Munich, Germany

24 Munich Cluster for Systems Neurology (SyNergy) Munich, Munich, Germany

25 Ageing Epidemiology Research Unit (AGE), School of Public Health, Imperial College London, London, UK

26 Institute for Clinical Radiology, Ludwig-Maximilians-University, Marchioninistr. 15, 81377 Munich

27 MR-Research in Neurosciences, Department of Cognitive Neurology, Georg-August-University Goettingen, Germany

28 Bernstein Center for Computational Neuroscience, Charité — Universitätsmedizin, Berlin, Germany

29 Department for Biomedical Magnetic Resonance, University of Tübingen, 72076 Tübingen, Germany

30 Study Center Bonn, Medical Faculty, Venusberg-Campus 1, 53127 Bonn, Germany

31 Department of Medical Imaging, University of Toronto, Toronto, Canada

32 Department of Nuclear Medicine, Charité - Universitätsmedizin Berlin, corporate member of Freie Universität Berlin, Humboldt-Universität zu Berlin, and Berlin Institute of Health, Berlin, Germany

33 Department of Diagnostic and Interventional Radiology and Nuclear Medicine, University Medical Center Hamburg-Eppendorf, Hamburg, Germany

34 Department of Nuclear Medicine, University Hospital Bonn, Bonn, Germany

35 Department of Nuclear Medicine, Faculty of Medicine and University Hospital Cologne, University of Cologne, Germany

36 Department of Nuclear Medicine, Ludwig-Maximilian-University Munich, Munich, Germany

37 Department of Nuclear Medicine, Rostock University Medical Centre, Rostock

38 Department of Nuclear Medicine and Clinical Molecular Imaging, Eberhard-Karls-University, Tuebingen, Germany

39 Excellence Cluster on Cellular Stress Responses in Aging-Associated Diseases (CECAD), University of Cologne, Joseph-Stelzmann-Strasse 26, 50931Köln, Germany

40 Neurosciences and Signaling Group, Institute of Biomedicine (iBiMED), Department of Medical Sciences, University of Aveiro, Aveiro, Portugal

41 Division of Neurogenetics and Molecular Psychiatry, Department of Psychiatry and Psychotherapy, Faculty of Medicine and University Hospital Cologne, University of Cologne, Cologne, Germany

42 Institute for Medical Biometry, Informatics and Epidemiology, University Hospital Bonn, Venusberg-Campus 1, D-53127 Bonn
43 Department of Neuroradiology, University Hospital Bonn, Bonn, Germany
44 Clininical Functional Imaging Group, Department of Diagnostic and Interventional Radiology, University Hospital Bonn, Bonn, Germany
45 School of Medicine, Technical University of Munich; Department of Psychiatry and Psychotherapy, Munich, Germany
46 University of Edinburgh and UK DRI, Edinburgh, UK
47 Department of Psychiatry & Glenn Biggs Institute for Alzheimer's and Neurodegenerative Diseases, San Antonio, TX, USA
48 Glenn Biggs Institute for Alzheimer's and Neurodegenerative Diseases, UT Health San Antonio, San Antonio, Texas, USA.
49 Berlin Center for Advanced Neuroimaging, Charité – Universitätsmedizin Berlin, Berlin, Germany
50 Department of Psychiatry and Psychotherapy, University of Tübingen, Tübingen, Germany
51 Department of Nuclear Medicine, University Hospital Bonn, Bonn, Germany
52 Sheffield Institute for Translational Neuroscience (SITraN), University of Sheffield, Sheffield, UK
53 Department of Neuroradiology, University Hospital LMU, Munich, Germany
54 Institute of Neuroscience and Medicine (INM-2), Molecular Organization of the Brain, Forschungszentrum Jülich, Germany
55 Department of Nuclear Medicine, Inselspital, Bern University Hospital, University of Bern, Switzerland
56 Department for Psychiatry and Psychotherapy, University Clinic Magdeburg, Magdeburg, Germany
57 Luxembourg Centre for Systems Biomedicine (LCSB), University of Luxembourg, L-4367 Belvaux
58 Charité Universitätsmedizin Berlin, Department of Psychiatry and Neurosciences, Campus Benjamin Franklin
59 German Center for Mental Health (DZPG), partner site Berlin
60 Leibniz Institute for Neurobiology, Brenneckestr. 6, 39118, Magdeburg, Germany
61 Charité – Universitätsmedizin Berlin, ECRC Experimental and Clinical Research Center, Berlin, Germany
62 Department of Neurology, XuanWu Hospital of Capital Medical University, Changchun Street 45, 100053, Beijing, China
63 Division of Translational Genomics of Neurodegenerative Diseases, Hertie Institute for Clinical Brain Research and Center of Neurology, University of Tübingen, Tübingen, Germany
64 Department of Psychiatry and Psychotherapy, School of Medicine and Health, Technical University of Munich, and German Center for Mental Health (DZPG), Munich, Germany

### Supplementary methods

#### DELCODE inclusion criteria

DELCODE comprises two control groups and three patient groups. Clinical screening of patient groups (subjective cognitive decline [SCD], mild cognitive impairment [MCI], and dementia of the Alzheimer type [DAT]) was performed at the respective memory clinic. Those in the SCD group fulfilled research criteria for SCD, expressed concerns about a subjectively perceived cognitive decline to a physician at the respective memory clinic, and were cognitively unimpaired as defined by a score better than −1.5 *SD* below the German age, sex, and education-adjusted norms for the Consortium to Establish a Registry for Alzheimer's Disease (CERAD) test battery. The MCI group fulfilled research criteria for MCI and was limited to amnestic MCI as defined by a performance below −1.5 *SD* on the delayed recall trial of the CERAD word-list episodic memory tests. The DAT group fulfilled research criteria for DAT and had Mini-Mental Status Examination scores ≥ 18. Both control groups included cognitively unimpaired individuals with one group specifically including first-degree relatives of AD patients ($n$ = 56). The other control group comprised $n$ = 139 participants. The two control groups were collapsed to form the HC group in the present study. Participants in this diagnostic group performed better than −1.5 *SD* below the German age, sex, and education-adjusted norms for the CERAD test battery and reported no subjective concerns about cognitive decline.

For all diagnostic groups, inclusion criteria were age ≥ 60 years, fluency in German, the ability to provide informed consent, and the availability of a study partner. Exclusion criteria included, but were not limited to, conditions that would prevent participation according to the study protocol (e.g., significant sensorimotor impairments), current or past diagnoses of psychiatric disorders (e.g., major depressive disorder, disorders due to psychoactive substance use), neurodegenerative disorders other than AD, vascular dementia, and a history of stroke with persistent clinical symptoms. The use of sedative, anticholinergic, and other anti-dementia medication as well as the use of investigational agents for treatment of cognitive impairment or dementia in all diagnostic groups except for the DAT group up to ≤ 1 month prior to screening and during the expected duration of the study led to exclusion.

#### CSF amyloid positivity predictions

We closely followed an approach proposed by Hu *et al.* to predict cerebrospinal fluid (CSF)-based positivity for amyloid-β (Aβ) in individuals without CSF data.[1] This analysis was performed in the whole DELCODE dataset and was not limited to sub-sample analysed in the present manuscript. First, we fitted a binomial logistic regression model predicting CSF-based Aβ-positivity in participants with available CSF data and all required predictors (number of apolipoprotein E [APOE] ε4 alleles [0/1/2], APOE ε2 carriership [yes/no], age, plasma $A\beta_{42}/A\beta_{40}$ ratio, and plasma phosphorylated tau 181). These data availability criteria resulted in a sample size of $n$ = 328. Note that the original work by Hu *et al.* did not include plasma phosphorylated tau 181 as a predictor. We included this metric as this led to a substantial increase in model performance (AUC [95% confidence interval]: 0.94 [0.91; 1.00] *versus* 0.91 [0.87; 1.00]). The trained model was then used to predict the probability of CSF-based Aβ-positivity in all DELCODE participants with available predictors ($n$ = 675), regardless of CSF biomarker availability. The extracted subject-level probabilities ranged from 0 to 1, with higher values representing a higher probability of an individual being positive based on CSF $A\beta_{42}/A\beta_{40}$ ratios.

#### Longitudinal segmentation of medial temporal lobe subregions

The longitudinal, template-based implementation of the ASHS algorithm[2] relies on within-subject multivariate templates that were created using Advanced Normalization Tools (ANTs, ref.[3]). For T1-ASHS, these templates were calculated from all available T1-weighted images for a given participant, both at the original and an upsampled (0.5 × 0.5 × 1.0mm) resolution to approximate the anisotropic resolution of the T2-weighted images.[4] For T2-ASHS, templates were generated from available T1- and T2-weighted images. ASHS was run on the resulting within-subject multivariate templates, using the atlases referenced in the main text. To obtain segmentations for each available time point, the resulting multilabel masks were warped onto the individual upsampled T1-weighted (for T1-ASHS) or original T2-weighted (for T2-ASHS) images using the warp fields and transformation matrices created during template calculation. All template segmentations underwent visual quality control and were manually edited if needed.

### Supplementary results

#### Pre-normalization of cognitive test scores

Using latent process modelling with subject-level random slopes and intercepts as well as spline-based link functions, we pre-normalized raw scores on the ADAS-Cog Delayed Word Recall (five knots with internal nodes at 2, 5, and 7), ADAS-Cog Figure Copying (four knots with internal nodes at 7 and 9), ADAS-Cog-13 (four quantile based knots), CDR-SB (six knots with internal nodes at 2, 5, 7, and 10), Clock Copying (five knots with internal nodes at 5, 7, and 9), Clock Drawing (five knots with internal nodes at 5, 7, and 9), FAQ (six knots with internal nodes at 2, 5, 10, 15), MMSE (six knots with internal nodes at 20, 24, 26, 28), and NPI-Q (six knots with internal nodes at 2, 5, 10, 15). This was done to ease LGCM fitting given the non-normal distributions (Supplementary Figure 9). These scores, along with the non-normalized raw scores from the remaining inventories, were then *z*-scored to participants in the HC Aβ–, HC Aβ+, and SCD Aβ+ groups and then passed latent growth curve models (LGCMs). These data are displayed in Supplementary Figure 10.

#### Latent growth curve model fits

None of the fitted LGCMs failed to meet all of the *a priori* defined criteria for acceptable model fit. Only three models slightly surpassed the threshold of RMSEA = 0.08 (hippocampal tail & PACC-5: RMSEA = 0.082; hippocampal tail & FCSRT-96: RMSEA = 0.080; inferior parietal cortex & PACC-5: RMSEA = 0.082). Given their acceptable CFIs (all > 0.96), these models were retained but should be interpreted cautiously.
With regards to variance structure, parahippocampal volumetric slopes did not consistently meet the critical *Z* threshold in all cases. However, as these violations were marginal and some models including this measure did show sufficient slope variance, we chose to include all models incorporating parahippocampal volumes, while advising readers to consider this limitation.

### Supplementary figures

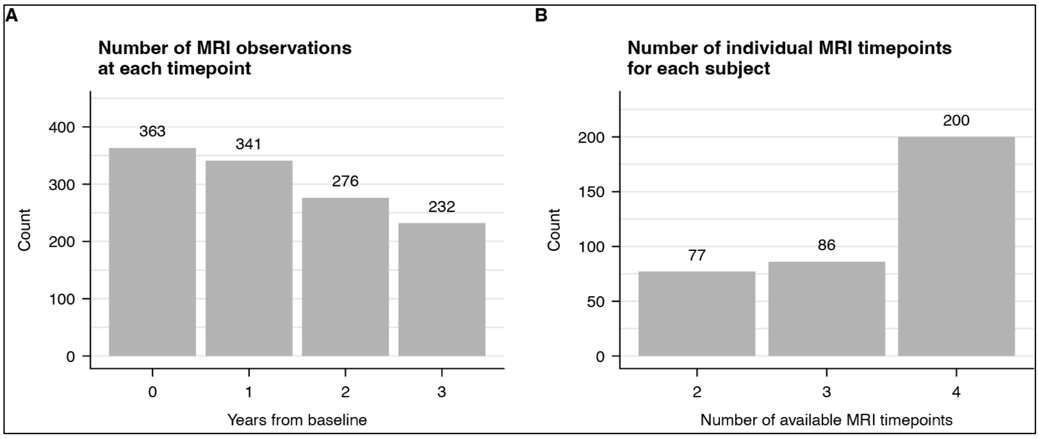


**Supplementary Figure 1 MRI data availability.** Bar plots indicate (**A**) the number of available scans by study time point and (**B**) the number of subjects with two, three, or four available scans.

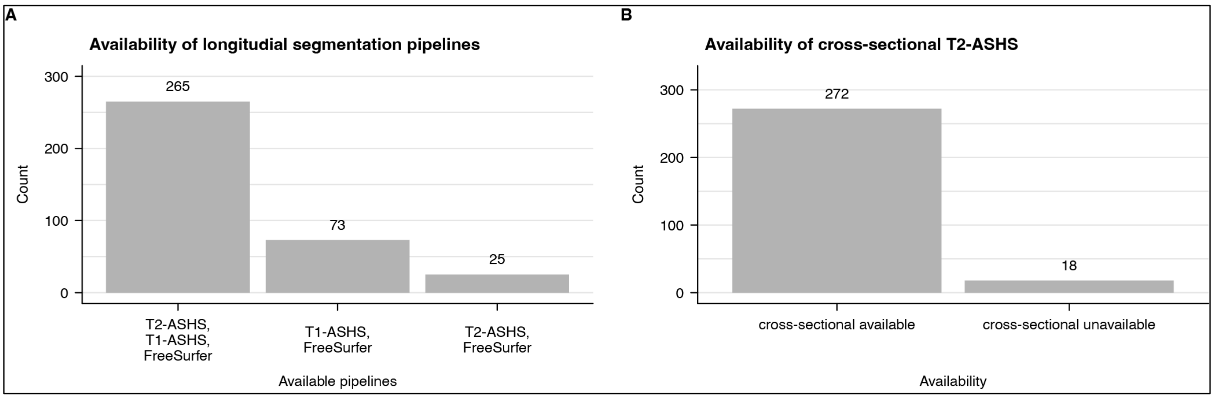


**Supplementary Figure 2 Segmentation pipeline availability.** Displayed are the counts of (**A**) different constellations of segmentation pipeline availability and (**B**) the availability of cross-sectional T2-ASHS in participants with available longitudinal T2-ASHS. The availability of either T1- or T2-ASHS were required for inclusion.

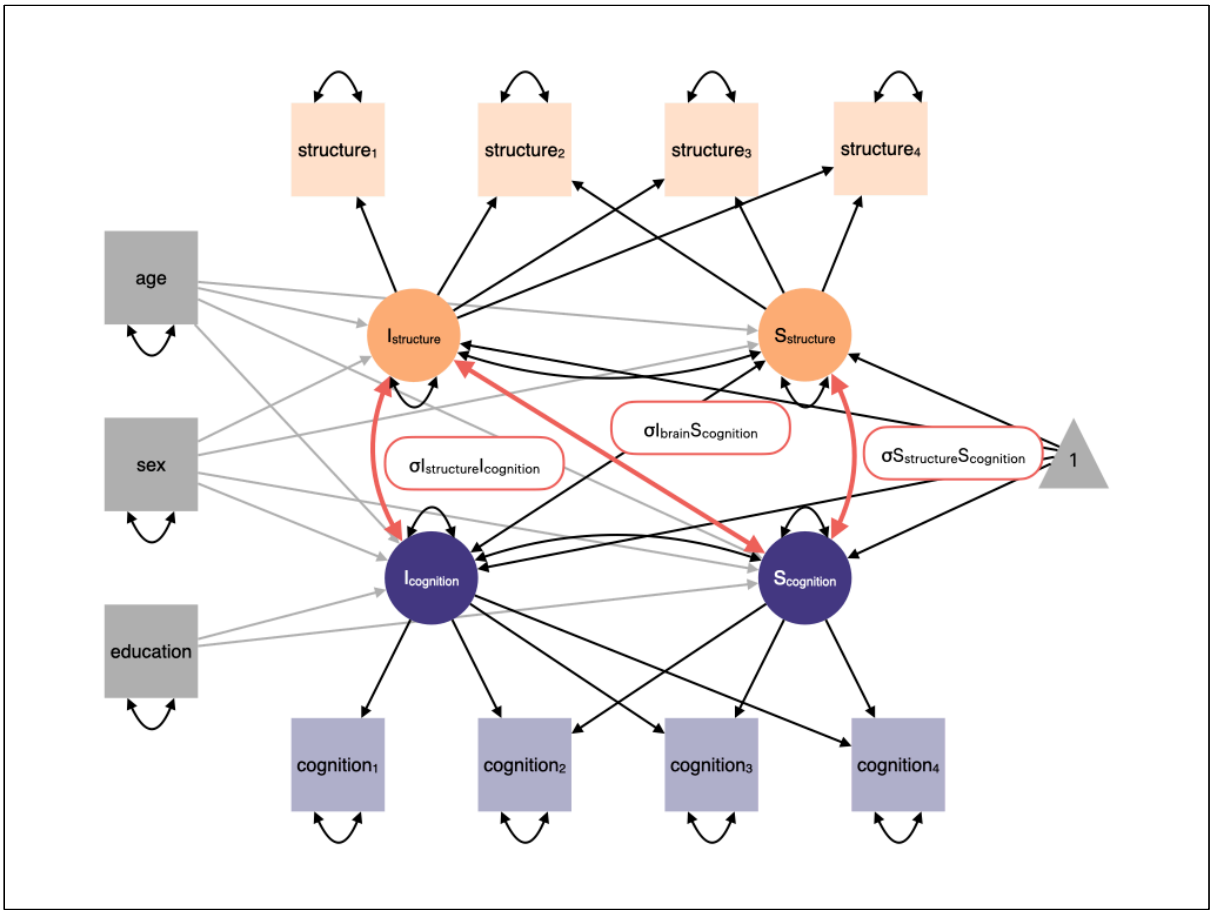


**Supplementary Figure 3 Bivariate latent growth curve model (LGCM) estimating the associations of longitudinally recorded measures of brain structure and cognition.** Manifest variables were recorded once (covariates) or four times (structure and cognitive variables). Parameters of interest were the covariance estimates representing baseline-baseline ($\sigma I_{structure} I_{cognition}$), baseline-change ($\sigma I_{structure} S_{cognition}$), and change-change ($\sigma S_{structure} S_{cognition}$) associations of structure and cognition. Paths estimating the residual covariance between observed cognitive and brain variables at each time point are not shown for visual clarity.

Abbreviations: I, intercept; S, slope.

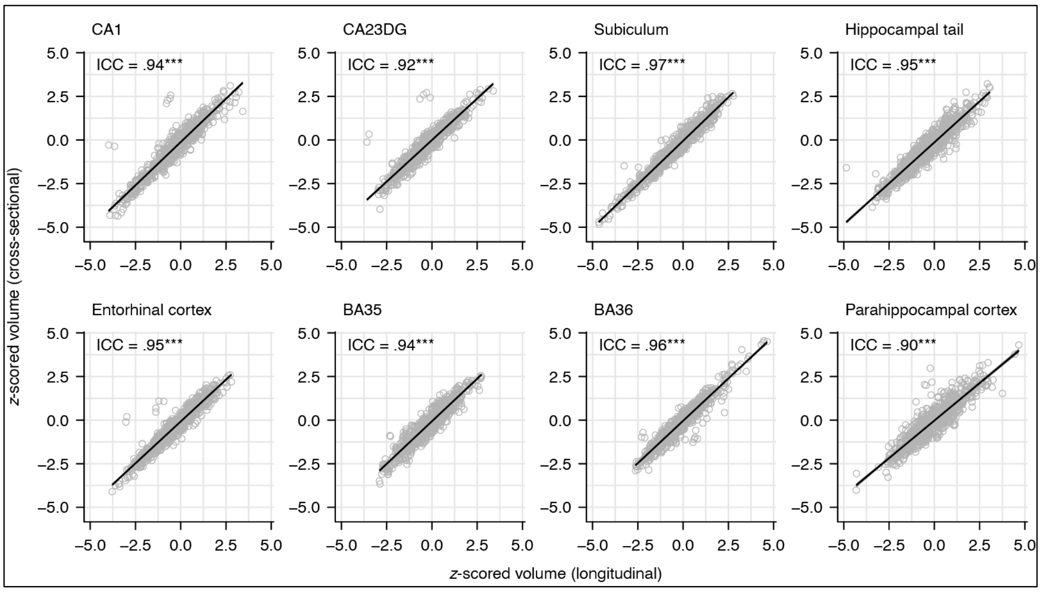


**Supplementary Figure 4 Scatter plots of baseline volumes obtained from the template-based and cross-sectional implementations of T2-ASHS.** Volumes were scaled and centred to the HC Aβ– group.
Abbreviations: ICC, intraclass correlation coefficient.

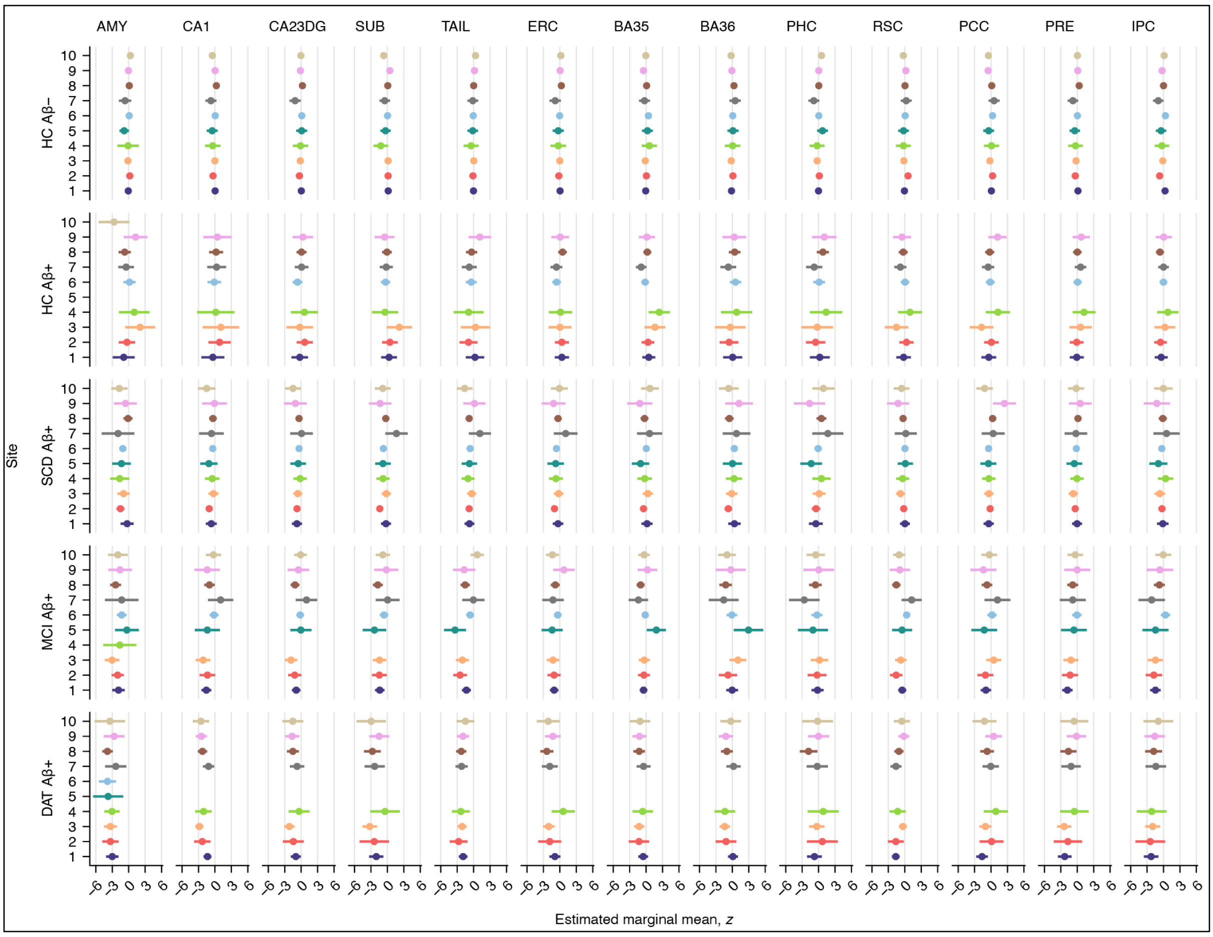


**Supplementary Figure 5 Estimated marginal means of the analysed structural MRI readouts, stratified by diagnostic group and DELCODE scanning site.** All readouts were scaled and centred to the HC Aβ– group. Estimated marginal means are controlled for age and sex. Error bars denote 95% confidence intervals. Abbreviations: Aβ, amyloid-β. AMY, amygdala. BA, Brodmann area. CA, cornu ammonis. CA23DG, cornu ammonis 2, 3, and dentate gyrus. DAT, dementia of the Alzheimer type. ERC, entorhinal cortex. FDR, false discovery rate. HC, healthy controls. IPC, inferior parietal cortex. MCI, mild cognitive impairment. PCC, posterior cingulate cortex. PHC, parahippocampal cortex. PRE, precuneus. RSC, retrosplenial cortex. SCD, subjective cognitive decline. SUB, subiculum. TAIL, hippocampal tail.

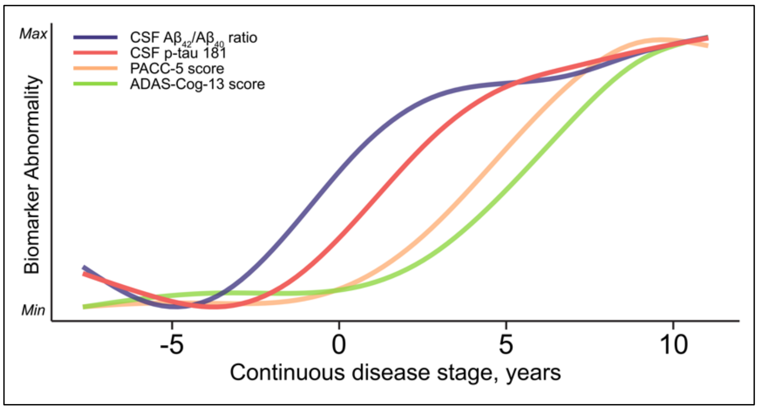


**Supplementary Figure 6 Biomarker trajectories across a continuous disease stage.** This model was modified from Lattmann-Grefe *et al.* by excluding structural MRI markers in order to prevent circular hypothesis testing in the current study.

Abbreviations: ADAS-Cog, Alzheimer's Disease Assessment Scale-Cognitive subscale. PACC-5, Preclinical Alzheimer Cognitive Composite.

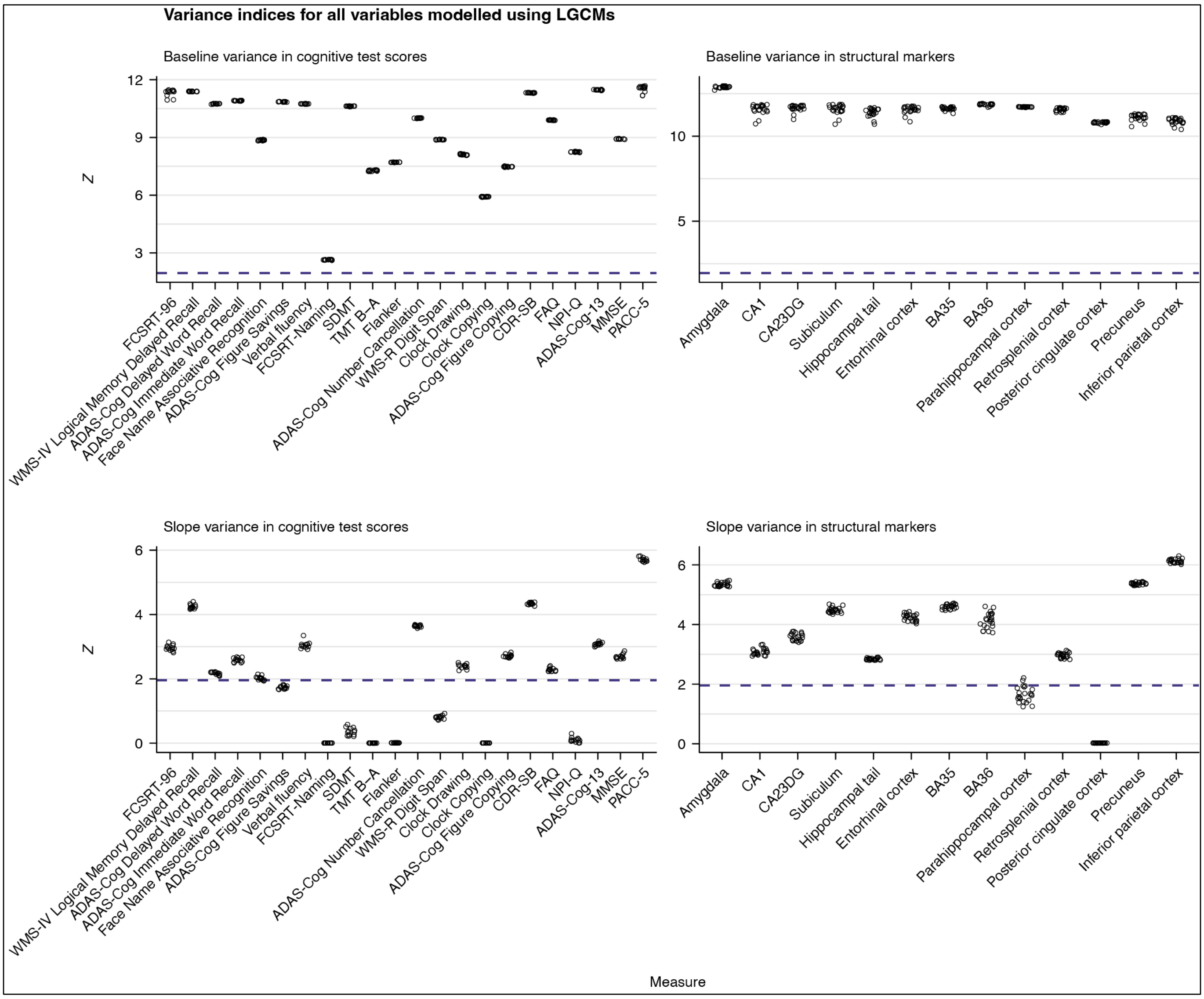


**Supplementary Figure 7 Variance indices for all structural markers and cognitive test scores included in the LGCM analysis.** The critical $Z = 1.96$ threshold is indicated by the dashed line. Measures falling below that threshold do not have sufficient variance to allow inference about covariance with other variables.

Abbreviations: ADAS-Cog, Alzheimer's Disease Assessment Scale-Cognitive subscale. BA, Brodmann area. CA, cornu ammonis. CA23DG, cornu ammonis 2, 3, and dentate gyrus. CSD-SB, Clinical Dementia Rating Sum of Boxes scale. FAQ, Functional Activities Questionnaire. FCSRT, Free and Cued Selective Reminding Test. MMSE, Mini-Mental State Examination. NPI-Q, Neuropsychiatric Inventory–Questionnaire. PACC-5, Preclinical Alzheimer Cognitive Composite. TMT, Trail Making Test. WMS, Wechsler Memory Scale.

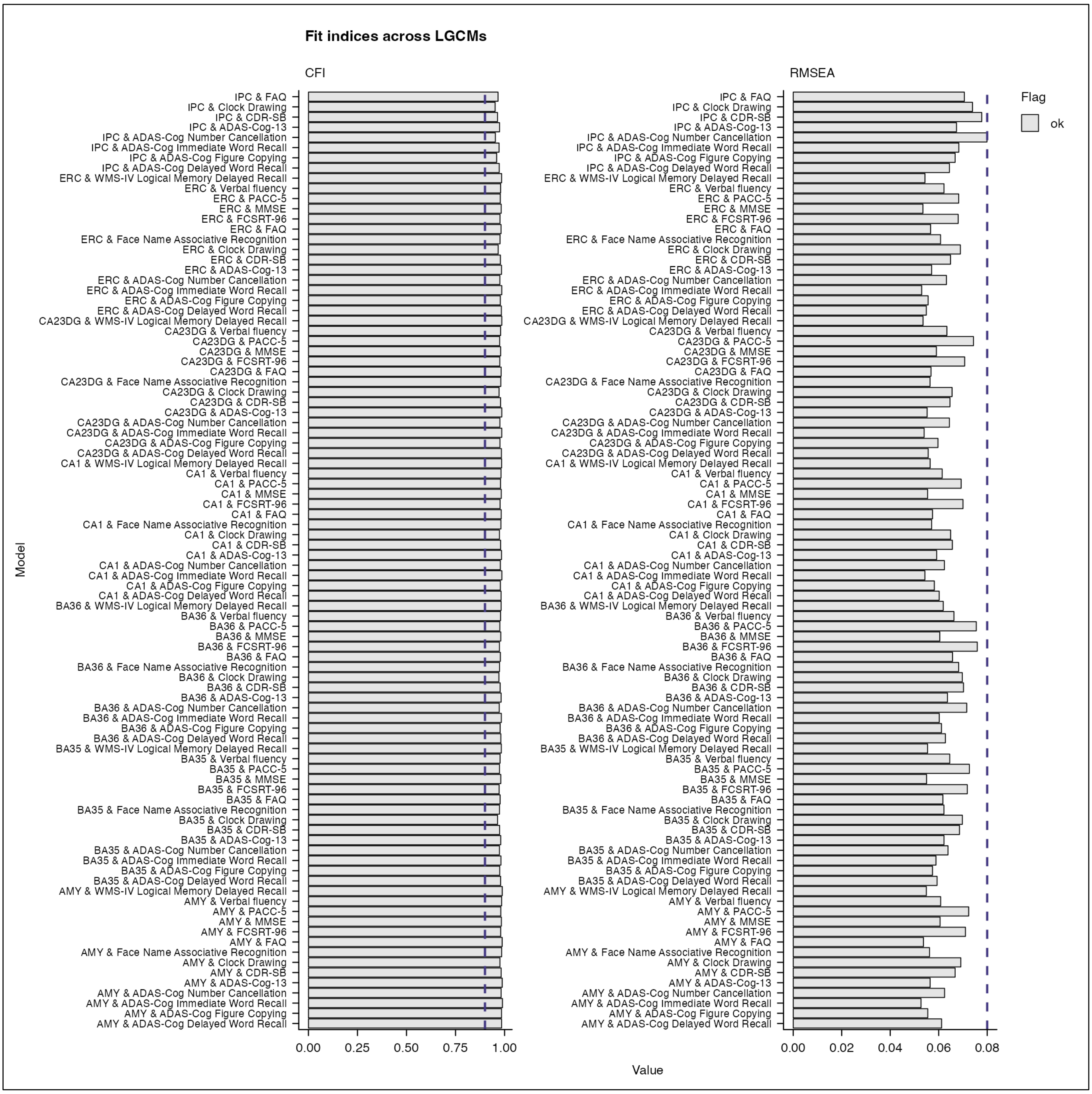


**Supplementary Figure 8 Fit indices for all LGCMs with sufficient variance in slopes and intercepts of both cognitive test scores and structural MRI markers.** The critical values of RMSEA = 0.08 and CFI = 0.90 are indicated by the dashed lines.

Abbreviations: ADAS-Cog, Alzheimer's Disease Assessment Scale-Cognitive subscale. BA, Brodmann area. CA, cornu ammonis. CA23DG, cornu ammonis 2, 3, and dentate gyrus. CFI, comparative fit index. CSD-SB, Clinical Dementia Rating Sum of Boxes scale. FAQ, Functional Activities Questionnaire. FCSRT, Free and Cued Selective Reminding Test. MMSE, Mini-Mental State Examination. PACC-5, Preclinical Alzheimer Cognitive Composite. RMSEA, root mean square error of approximation.

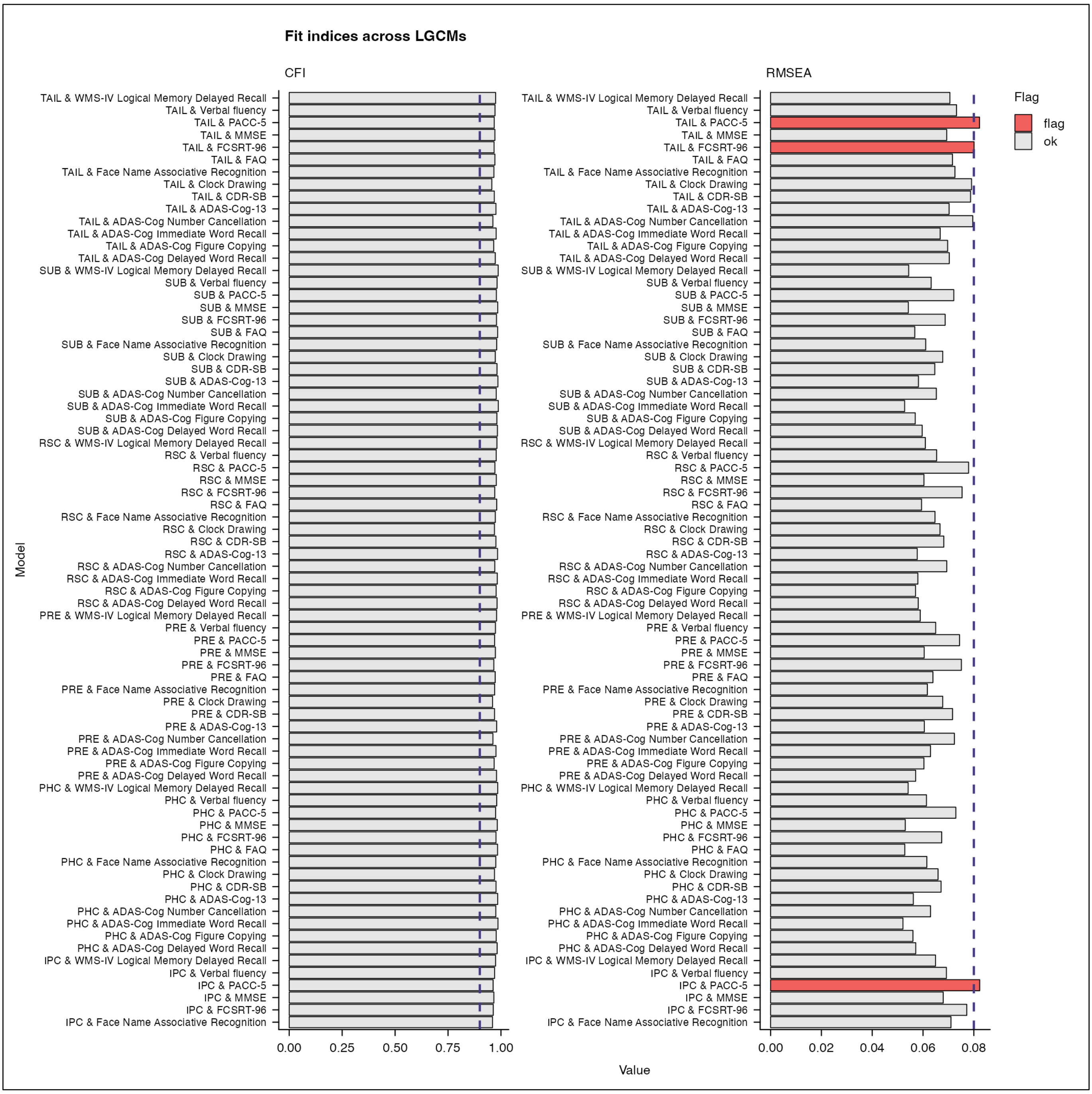


**Supplementary Figure 8** *continued*

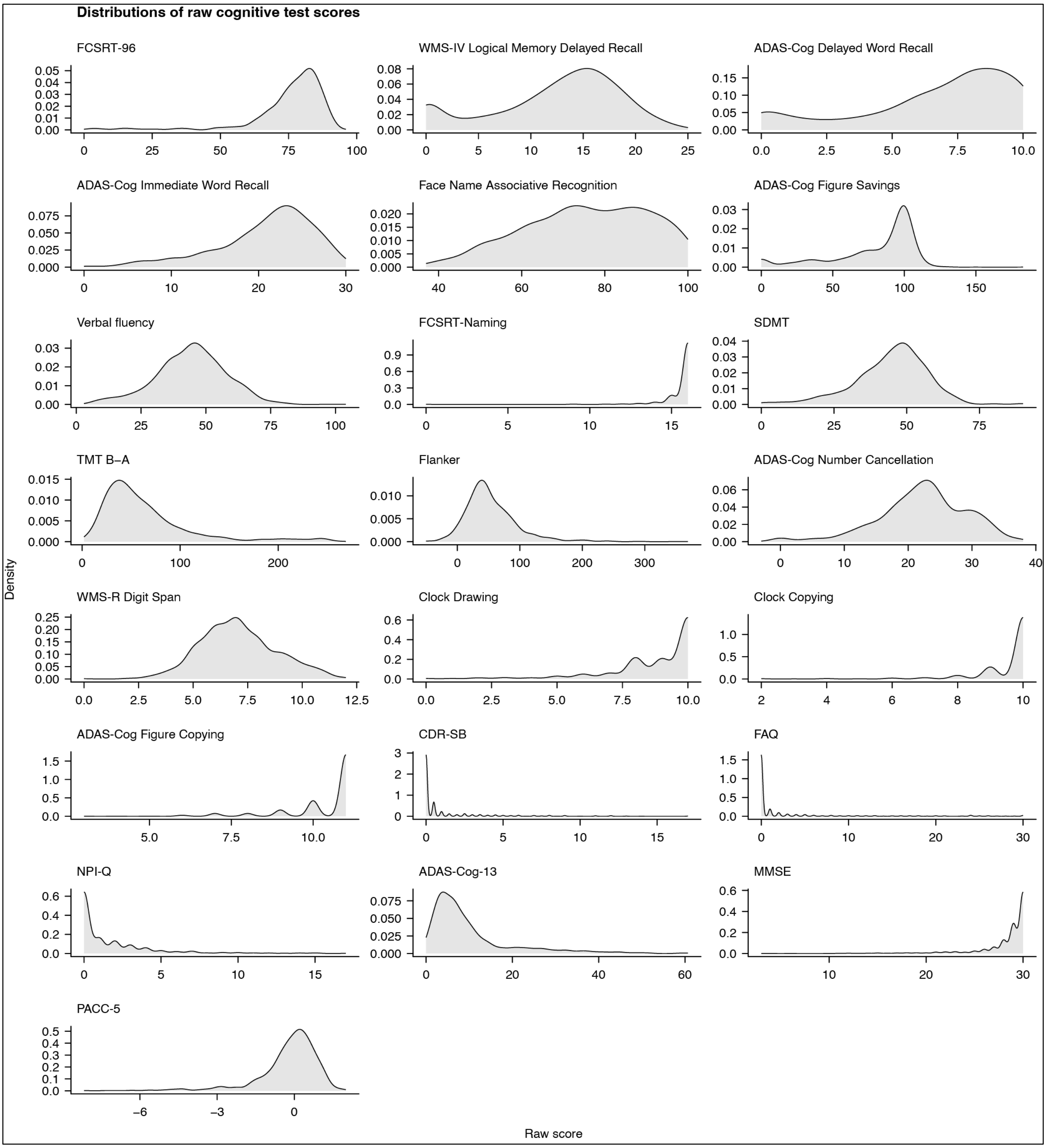


**Supplementary Figure 9 Density plots of all raw cognitive test scores.**

Abbreviations: ADAS-Cog, Alzheimer's Disease Assessment Scale-Cognitive subscale. CSD-SB, Clinical Dementia Rating Sum of Boxes scale. FAQ, Functional Activities Questionnaire. FCSRT, Free and Cued Selective Reminding Test. MMSE, Mini-Mental State Examination. NPI-Q, Neuropsychiatric Inventory–Questionnaire. PACC-5, Preclinical Alzheimer Cognitive Composite. SDMT, Symbol-Digit Modalities Test. TMT, trail making test. WMS, Wechsler Memory Scale.

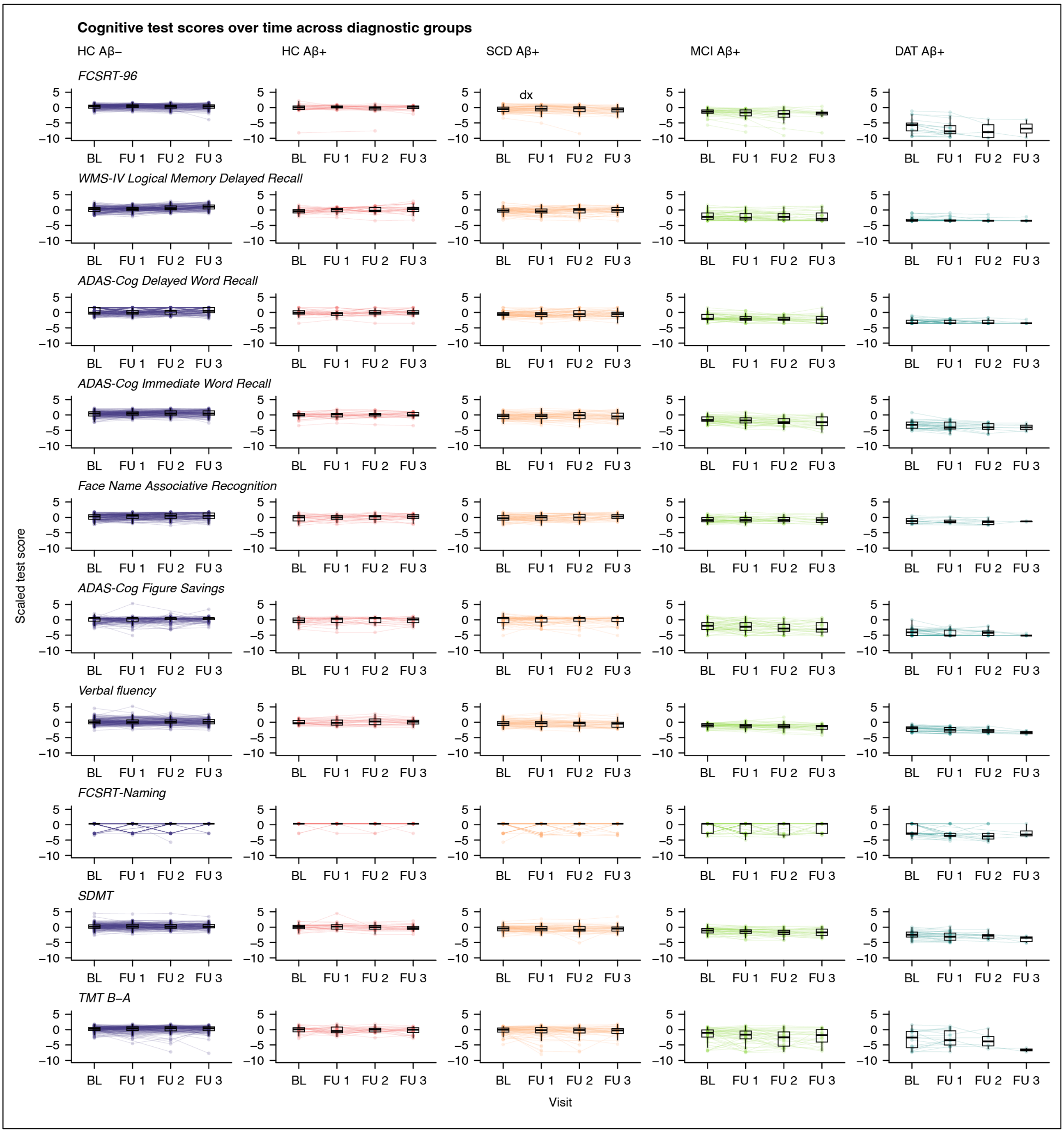


**Supplementary Figure 10 Standardized cognitive test scores by inventory, diagnostic group, and visit.**

Abbreviations: ADAS-Cog, Alzheimer's Disease Assessment Scale-Cognitive subscale. BL, baseline. CSD-SB, Clinical Dementia Rating Sum of Boxes scale. DAT, dementia of the Alzheimer type. FAQ, Functional Activities Questionnaire. FCSRT, Free and Cued Selective Reminding Test. FU, follow-up. HC, healthy controls. MCI, mild cognitive impairment. MMSE, Mini-Mental State Examination. NPI-Q, Neuropsychiatric Inventory–Questionnaire. PACC-5, Preclinical Alzheimer Cognitive Composite. SCD, subjective cognitive decline. SDMT, Symbol-Digit Modalities Test. TMT, trail making test. WMS, Wechsler Memory Scale.

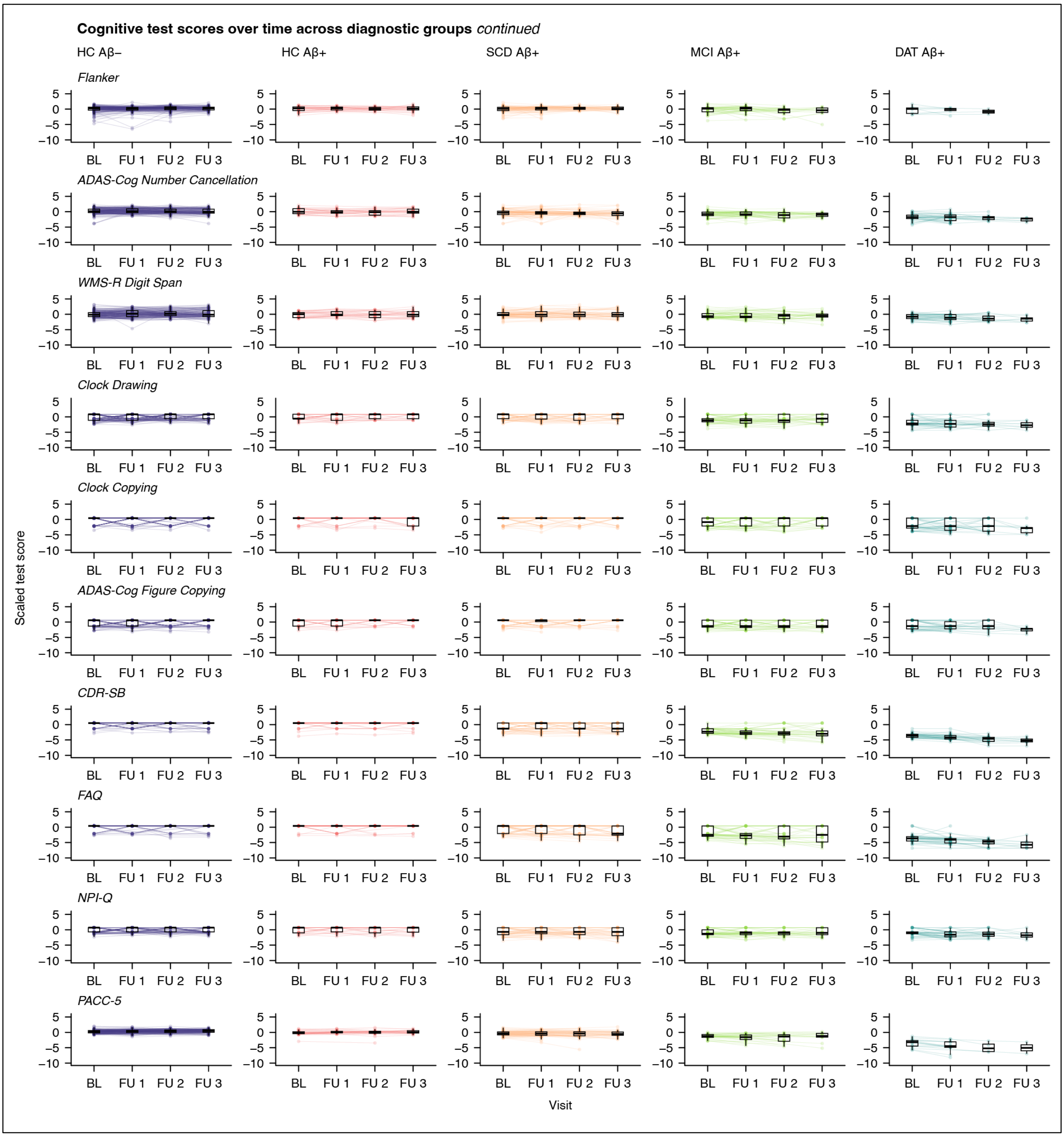


**Supplementary Figure 10** *continued*

### Supplementary tables

**Supplementary Table 1** Summary statistics for ANCOVA models estimating the effect of study site on scaled structural MRI readouts while controlling for participant age and sex, stratified by diagnostic group.

| Marker | Diagnostic group | df | F | $p_{FDR}$ |
|---|---|---|---|---|
| Amygdala, volume | HC Aβ– | 9; 136 | 0.84 | .838 |
| | HC Aβ+ | 8; 16 | 1.58 | .894 |
| | SCD Aβ+ | 9; 64 | 1.12 | .924 |
| | MCI Aβ+ | 9; 38 | 0.99 | .551 |
| | DAT Aβ+ | 9; 25 | 0.46 | .953 |
| CA1, volume | HC Aβ– | 9; 139 | 0.97 | .765 |
| | HC Aβ+ | 7; 18 | 0.23 | .973 |
| | SCD Aβ+ | 9; 49 | 0.65 | .924 |
| | MCI Aβ+ | 8; 21 | 1.67 | .357 |
| | DAT Aβ+ | 7; 8 | 2.38 | .653 |
| CA23DG, volume | HC Aβ– | 9; 139 | 0.97 | .765 |
| | HC Aβ+ | 7; 18 | 0.55 | .894 |
| | SCD Aβ+ | 9; 49 | 0.48 | .924 |
| | MCI Aβ+ | 8; 21 | 1.85 | .352 |
| | DAT Aβ+ | 7; 8 | 1.02 | .792 |
| Subiculum, volume | HC Aβ– | 9; 139 | 1.51 | .765 |
| | HC Aβ+ | 7; 18 | 0.90 | .894 |
| | SCD Aβ+ | 9; 49 | 1.77 | .924 |
| | MCI Aβ+ | 8; 21 | 1.13 | .497 |
| | DAT Aβ+ | 7; 8 | 0.92 | .792 |
| Hippocampal tail, volume | HC Aβ– | 9; 139 | 0.28 | .980 |
| | HC Aβ+ | 7; 18 | 0.59 | .894 |
| | SCD Aβ+ | 9; 49 | 0.82 | .924 |
| | MCI Aβ+ | 8; 21 | 3.02 | .131 |
| | DAT Aβ+ | 7; 8 | 0.31 | .953 |
| Entorhinal cortex, volume | HC Aβ– | 9; 139 | 0.66 | .842 |
| | HC Aβ+ | 7; 18 | 0.85 | .894 |
| | SCD Aβ+ | 9; 49 | 0.71 | .924 |
| | MCI Aβ+ | 8; 21 | 0.93 | .553 |
| | DAT Aβ+ | 7; 8 | 1.64 | .653 |
| BA35, volume | HC Aβ– | 9; 139 | 1.11 | .765 |
| | HC Aβ+ | 7; 18 | 2.13 | .894 |
| | SCD Aβ+ | 9; 49 | 0.72 | .924 |
| | MCI Aβ+ | 8; 21 | 1.55 | .371 |
| | DAT Aβ+ | 7; 8 | 0.50 | .953 |
| BA36, volume | HC Aβ– | 9; 139 | 0.62 | .842 |
| | HC Aβ+ | 7; 18 | 0.58 | .894 |
| | SCD Aβ+ | 9; 49 | 1.10 | .924 |
| | MCI Aβ+ | 8; 21 | 1.79 | .352 |
| | DAT Aβ+ | 7; 8 | 1.76 | .653 |
| Parahippocampal cortex, volume | HC Aβ– | 9; 139 | 1.12 | .765 |
| | HC Aβ+ | 7; 18 | 0.76 | .894 |
| | SCD Aβ+ | 9; 49 | 1.01 | .924 |
| | MCI Aβ+ | 8; 21 | 0.50 | .843 |
| | DAT Aβ+ | 7; 8 | 0.90 | .792 |
| Retrosplenial cortex, average thickness | HC Aβ– | 9; 139 | 0.71 | .842 |
| | HC Aβ+ | 7; 18 | 0.75 | .894 |
| | SCD Aβ+ | 9; 49 | 0.59 | .924 |
| | MCI Aβ+ | 8; 21 | 3.32 | .131 |
| | DAT Aβ+ | 7; 8 | 2.20 | .653 |
| Posterior cingulate cortex, average thickness | HC Aβ– | 9; 139 | 1.32 | .765 |
| | HC Aβ+ | 7; 18 | 1.13 | .894 |
| | SCD Aβ+ | 9; 49 | 1.33 | .924 |
| | MCI Aβ+ | 8; 21 | 1.13 | .497 |
| | DAT Aβ+ | 7; 8 | 1.73 | .653 |
| Precuneus, average thickness | HC Aβ– | 9; 139 | 1.22 | .765 |
| | HC Aβ+ | 7; 18 | 0.50 | .894 |
| | SCD Aβ+ | 9; 49 | 0.47 | .924 |
| | MCI Aβ+ | 8; 21 | 1.25 | .497 |
| | DAT Aβ+ | 7; 8 | 1.38 | .714 |
| Inferior parietal cortex, average thickness | HC Aβ– | 9; 139 | 2.14 | .387 |
| | HC Aβ+ | 7; 18 | 0.60 | .894 |
| | SCD Aβ+ | 9; 49 | 0.41 | .924 |
| | MCI Aβ+ | 8; 21 | 1.98 | .352 |
| | DAT Aβ+ | 7; 8 | 0.26 | .953 |

FDR correction was performed separately in each diagnostic groups across structural measures.
Abbreviations: DAT, dementia of the Alzheimer type. df, degrees of freedom. HC, healthy control. MCI, mild cognitive impairment. SCD, subjective cognitive decline.

**Supplementary Table 2** Summary statistics of ANCOVA models comparing baseline structural MRI readouts.

| Marker | Predictor | *SS* | *df* | *MS* | *F* | $\eta^2_{partial}$ [95% C.I.] | *p* |
|---|---|---|---|---|---|---|---|
| Amygdala, volume | Age, years | 169.25 | 1; 331 | 169.25 | 111.34 | 0.25 [0.19; 1.00] | < .001*** |
| | Sex, male | 0.59 | 1; 331 | 0.59 | 0.39 | 0.00 [0.00; 1.00] | .533 |
| | Diagnostic group | 271.98 | 4; 331 | 67.99 | 44.73 | 0.35 [0.28; 1.00] | < .001*** |
| CA1, volume | Age, years | 38.68 | 1; 283 | 38.68 | 34.81 | 0.11 [0.06; 1.00] | < .001*** |
| | Sex, male | 0.07 | 1; 283 | 0.07 | 0.07 | 0.00 [0.00; 1.00] | .798 |
| | Diagnostic group | 71.70 | 4; 283 | 17.92 | 16.13 | 0.19 [0.12; 1.00] | < .001*** |
| CA23DG, volume | Age, years | 21.94 | 1; 283 | 21.94 | 22.82 | 0.07 [0.03; 1.00] | < .001*** |
| | Sex, male | 2.08 | 1; 283 | 2.08 | 2.16 | 0.01 [0.00; 1.00] | .143 |
| | Diagnostic group | 28.24 | 4; 283 | 7.06 | 7.34 | 0.09 [0.04; 1.00] | < .001*** |
| Subiculum, volume | Age, years | 76.66 | 1; 283 | 76.66 | 76.48 | 0.21 [0.15; 1.00] | < .001*** |
| | Sex, male | 2.70 | 1; 283 | 2.70 | 2.69 | 0.01 [0.00; 1.00] | .102 |
| | Diagnostic group | 80.66 | 4; 283 | 20.16 | 20.11 | 0.22 [0.15; 1.00] | < .001*** |
| Hippocampal tail, volume | Age, years | 84.65 | 1; 283 | 84.65 | 87.17 | 0.24 [0.17; 1.00] | < .001*** |
| | Sex, male | 0.10 | 1; 283 | 0.10 | 0.10 | 0.00 [0.00; 1.00] | .747 |
| | Diagnostic group | 60.06 | 4; 283 | 15.01 | 15.46 | 0.18 [0.11; 1.00] | < .001*** |
| Entorhinal cortex, volume | Age, years | 27.88 | 1; 283 | 27.88 | 28.78 | 0.09 [0.05; 1.00] | < .001*** |
| | Sex, male | 0.27 | 1; 283 | 0.27 | 0.28 | 0.00 [0.00; 1.00] | .595 |
| | Diagnostic group | 40.25 | 4; 283 | 10.06 | 10.39 | 0.13 [0.07; 1.00] | < .001*** |
| BA35, volume | Age, years | 15.53 | 1; 283 | 15.53 | 16.20 | 0.05 [0.02; 1.00] | < .001*** |
| | Sex, male | 2.19 | 1; 283 | 2.19 | 2.28 | 0.01 [0.00; 1.00] | .132 |
| | Diagnostic group | 12.84 | 4; 283 | 3.21 | 3.35 | 0.05 [0.01; 1.00] | .011* |
| BA36, volume | Age, years | 7.68 | 1; 283 | 7.68 | 6.26 | 0.02 [0.00; 1.00] | .013* |
| | Sex, male | 2.81 | 1; 283 | 2.81 | 2.29 | 0.01 [0.00; 1.00] | .131 |
| | Diagnostic group | 7.99 | 4; 283 | 2.00 | 1.63 | 0.02 [0.00; 1.00] | .167 |
| Parahippocampal cortex, volume | Age, years | 10.30 | 1; 283 | 10.30 | 7.92 | 0.03 [0.00; 1.00] | .005** |
| | Sex, male | 1.99 | 1; 283 | 1.99 | 1.53 | 0.01 [0.00; 1.00] | .217 |
| | Diagnostic group | 6.16 | 4; 283 | 1.54 | 1.19 | 0.02 [0.00; 1.00] | .317 |
| Retrosplenial cortex, average thickness | Age, years | 22.19 | 1; 283 | 22.19 | 23.49 | 0.08 [0.03; 1.00] | < .001*** |
| | Sex, male | 0.00 | 1; 283 | 0.00 | 0.00 | 0.00 [0.00; 1.00] | .966 |
| | Diagnostic group | 14.82 | 4; 283 | 3.71 | 3.92 | 0.05 [0.01; 1.00] | .004** |
| Posterior cingulate cortex, average thickness | Age, years | 9.04 | 1; 283 | 9.04 | 8.76 | 0.03 [0.01; 1.00] | .003** |
| | Sex, male | 0.81 | 1; 283 | 0.81 | 0.79 | 0.00 [0.00; 1.00] | .376 |
| | Diagnostic group | 5.56 | 4; 283 | 1.39 | 1.35 | 0.02 [0.00; 1.00] | .253 |
| Precuneus, average thickness | Age, years | 26.57 | 1; 283 | 26.57 | 26.60 | 0.09 [0.04; 1.00] | < .001*** |
| | Sex, male | 0.33 | 1; 283 | 0.33 | 0.33 | 0.00 [0.00; 1.00] | .566 |
| | Diagnostic group | 45.70 | 4; 283 | 11.43 | 11.44 | 0.14 [0.07; 1.00] | < .001*** |
| Inferior parietal cortex, average thickness | Age, years | 46.26 | 1; 283 | 46.26 | 43.77 | 0.13 [0.08; 1.00] | < .001*** |
| | Sex, male | 4.19 | 1; 283 | 4.19 | 3.97 | 0.01 [0.00; 1.00] | .047* |
| | Diagnostic group | 43.96 | 4; 283 | 10.99 | 10.40 | 0.13 [0.07; 1.00] | < .001*** |

Abbreviations: BA, Brodmann area. CA, cornu ammonis. CA23DG, cornu ammonis 2, 3, and dentate gyrus. df, degrees of freedom. MS, mean squares. SS, sum of squares.

**Supplementary Table 3** Summary statistics for planned *post hoc* diagnostic group comparisons of baseline structural MRI markers.

| Marker | Contrast | *b* [95% C.I.] | Sigma | *t* | $p_{raw}$ | $p_{FDR}$ |
|---|---|---|---|---|---|---|
| Amygdala, volume | HC Aβ+ – HC Aβ– | 0.07 [–0.62; 0.77] | 0.26 | 0.28 | .783 | .783 |
| | SCD Aβ+ – HC Aβ– | –0.70 [–1.19; –0.21] | 0.19 | –3.75 | < .001*** | < .001*** |
| | MCI Aβ+ – HC Aβ– | –1.57 [–2.13; –1.01] | 0.21 | –7.40 | < .001*** | < .001*** |
| | DAT Aβ+ – HC Aβ– | –2.95 [–3.59; –2.31] | 0.24 | –12.21 | < .001*** | < .001*** |
| | SCD Aβ+ – HC Aβ+ | –0.77 [–1.50; –0.04] | 0.28 | –2.78 | .006** | .007** |
| | MCI Aβ+ – SCD Aβ+ | –0.87 [–1.47; –0.28] | 0.23 | –3.86 | < .001*** | < .001*** |
| | DAT Aβ+ – MCI Aβ+ | –1.38 [–2.09; –0.67] | 0.27 | –5.16 | < .001*** | < .001*** |
| CA1, volume | HC Aβ+ – HC Aβ– | 0.36 [–0.23; 0.94] | 0.22 | 1.60 | .111 | .111 |
| | SCD Aβ+ – HC Aβ– | –0.37 [–0.82; 0.09] | 0.17 | –2.13 | .034* | .047* |
| | MCI Aβ+ – HC Aβ– | –0.81 [–1.37; –0.25] | 0.21 | –3.81 | < .001*** | < .001*** |
| | DAT Aβ+ – HC Aβ– | –1.84 [–2.55; –1.13] | 0.27 | –6.84 | < .001*** | < .001*** |
| | SCD Aβ+ – HC Aβ+ | –0.72 [–1.36; –0.08] | 0.24 | –2.99 | .003** | .005** |
| | MCI Aβ+ – SCD Aβ+ | –0.44 [–1.05; 0.17] | 0.23 | –1.91 | .057 | .067 |
| | DAT Aβ+ – MCI Aβ+ | –1.03 [–1.85; –0.21] | 0.31 | –3.32 | .001** | .002** |
| CA23DG, volume | HC Aβ+ – HC Aβ– | 0.00 [–0.54; 0.55] | 0.21 | 0.02 | .985 | .985 |
| | SCD Aβ+ – HC Aβ– | –0.39 [–0.81; 0.04] | 0.16 | –2.42 | .016* | .038* |
| | MCI Aβ+ – HC Aβ– | –0.59 [–1.11; –0.07] | 0.20 | –2.97 | .003** | .011* |
| | DAT Aβ+ – HC Aβ– | –1.18 [–1.84; –0.52] | 0.25 | –4.70 | < .001*** | < .001*** |
| | SCD Aβ+ – HC Aβ+ | –0.39 [–0.98; 0.20] | 0.22 | –1.74 | .083 | .116 |
| | MCI Aβ+ – SCD Aβ+ | –0.20 [–0.77; 0.37] | 0.22 | –0.93 | .353 | .412 |
| | DAT Aβ+ – MCI Aβ+ | –0.59 [–1.35; 0.17] | 0.29 | –2.04 | .042* | .074 |
| Subiculum, volume | HC Aβ+ – HC Aβ– | 0.17 [–0.39; 0.73] | 0.21 | 0.81 | .419 | .419 |
| | SCD Aβ+ – HC Aβ– | –0.43 [–0.86; 0.00] | 0.16 | –2.65 | .009** | .013* |
| | MCI Aβ+ – HC Aβ– | –0.91 [–1.44; –0.38] | 0.20 | –4.50 | < .001*** | < .001*** |
| | DAT Aβ+ – HC Aβ– | –2.03 [–2.71; –1.35] | 0.26 | –7.94 | < .001*** | < .001*** |
| | SCD Aβ+ – HC Aβ+ | –0.60 [–1.21; 0.00] | 0.23 | –2.63 | .009** | .013* |
| | MCI Aβ+ – SCD Aβ+ | –0.48 [–1.06; 0.10] | 0.22 | –2.17 | .031* | .036* |
| | DAT Aβ+ – MCI Aβ+ | –1.12 [–1.90; –0.34] | 0.30 | –3.80 | < .001*** | < .001*** |
| Hippocampal tail, volume | HC Aβ+ – HC Aβ– | –0.08 [–0.63; 0.47] | 0.21 | –0.38 | .703 | .703 |
| | SCD Aβ+ – HC Aβ– | –0.37 [–0.79; 0.06] | 0.16 | –2.30 | .022* | .031* |
| | MCI Aβ+ – HC Aβ– | –0.94 [–1.46; –0.41] | 0.20 | –4.72 | < .001*** | < .001*** |
| | DAT Aβ+ – HC Aβ– | –1.73 [–2.40; –1.07] | 0.25 | –6.90 | < .001*** | < .001*** |
| | SCD Aβ+ – HC Aβ+ | –0.29 [–0.89; 0.31] | 0.23 | –1.28 | .201 | .234 |
| | MCI Aβ+ – SCD Aβ+ | –0.57 [–1.14; 0.00] | 0.22 | –2.63 | .009** | .016* |
| | DAT Aβ+ – MCI Aβ+ | –0.80 [–1.56; –0.03] | 0.29 | –2.74 | .006** | .015* |
| Entorhinal cortex, volume | HC Aβ+ – HC Aβ– | 0.00 [–0.55; 0.55] | 0.21 | –0.01 | .995 | .995 |
| | SCD Aβ+ – HC Aβ– | –0.44 [–0.86; –0.02] | 0.16 | –2.75 | .006** | .015* |
| | MCI Aβ+ – HC Aβ– | –0.75 [–1.27; –0.22] | 0.20 | –3.76 | < .001*** | < .001*** |
| | DAT Aβ+ – HC Aβ– | –1.39 [–2.05; –0.72] | 0.25 | –5.52 | < .001*** | < .001*** |
| | SCD Aβ+ – HC Aβ+ | –0.44 [–1.03; 0.16] | 0.23 | –1.95 | .052 | .073 |
| | MCI Aβ+ – SCD Aβ+ | –0.30 [–0.88; 0.27] | 0.22 | –1.41 | .160 | .187 |
| | DAT Aβ+ – MCI Aβ+ | –0.64 [–1.41; 0.13] | 0.29 | –2.21 | .028* | .049* |
| BA35, volume | HC Aβ+ – HC Aβ– | 0.16 [–0.39; 0.70] | 0.21 | 0.77 | .441 | .618 |
| | SCD Aβ+ – HC Aβ– | 0.05 [–0.37; 0.48] | 0.16 | 0.34 | .731 | .731 |
| | MCI Aβ+ – HC Aβ– | –0.21 [–0.73; 0.31] | 0.20 | –1.07 | .287 | .502 |
| | DAT Aβ+ – HC Aβ– | –0.80 [–1.46; –0.14] | 0.25 | –3.18 | .002** | .011* |
| | SCD Aβ+ – HC Aβ+ | –0.10 [–0.70; 0.49] | 0.22 | –0.47 | .642 | .731 |
| | MCI Aβ+ – SCD Aβ+ | –0.27 [–0.83; 0.30] | 0.22 | –1.23 | .218 | .502 |
| | DAT Aβ+ – MCI Aβ+ | –0.58 [–1.35; 0.18] | 0.29 | –2.03 | .044* | .152 |
| Retrosplenial cortex, average thickness | HC Aβ+ – HC Aβ– | –0.12 [–0.67; 0.42] | 0.20 | –0.60 | .546 | .637 |
| | SCD Aβ+ – HC Aβ– | –0.12 [–0.54; 0.30] | 0.16 | –0.76 | .448 | .627 |
| | MCI Aβ+ – HC Aβ– | –0.45 [–0.97; 0.07] | 0.20 | –2.31 | .022* | .077 |
| | DAT Aβ+ – HC Aβ– | –0.88 [–1.54; –0.22] | 0.25 | –3.55 | < .001*** | .003** |
| | SCD Aβ+ – HC Aβ+ | 0.00 [–0.58; 0.59] | 0.22 | 0.01 | .988 | .988 |
| | MCI Aβ+ – SCD Aβ+ | –0.33 [–0.90; 0.23] | 0.21 | –1.55 | .122 | .238 |
| | DAT Aβ+ – MCI Aβ+ | –0.43 [–1.18; 0.33] | 0.29 | –1.50 | .136 | .238 |
| Precuneus, average thickness | HC Aβ+ – HC Aβ– | 0.40 [–0.15; 0.96] | 0.21 | 1.92 | .056 | .065 |
| | SCD Aβ+ – HC Aβ– | –0.08 [–0.51; 0.35] | 0.16 | –0.50 | .618 | .618 |
| | MCI Aβ+ – HC Aβ– | –0.62 [–1.15; –0.09] | 0.20 | –3.07 | .002** | .008** |
| | DAT Aβ+ – HC Aβ– | –1.40 [–2.07; –0.73] | 0.26 | –5.49 | < .001*** | < .001*** |
| | SCD Aβ+ – HC Aβ+ | –0.49 [–1.09; 0.12] | 0.23 | –2.12 | .035* | .048* |
| | MCI Aβ+ – SCD Aβ+ | –0.54 [–1.12; 0.04] | 0.22 | –2.44 | .015* | .027* |
| | DAT Aβ+ – MCI Aβ+ | –0.78 [–1.56; –0.01] | 0.29 | –2.66 | .008** | .019* |
| Inferior parietal cortex, average thickness | HC Aβ+ – HC Aβ– | 0.06 [–0.51; 0.63] | 0.22 | 0.28 | .776 | .776 |
| | SCD Aβ+ – HC Aβ– | –0.06 [–0.50; 0.38] | 0.17 | –0.35 | .728 | .776 |
| | MCI Aβ+ – HC Aβ– | –0.51 [–1.06; 0.04] | 0.21 | –2.45 | .015* | .035* |
| | DAT Aβ+ – HC Aβ– | –1.57 [–2.27; –0.88] | 0.26 | –6.00 | < .001*** | < .001*** |
| | SCD Aβ+ – HC Aβ+ | –0.12 [–0.74; 0.50] | 0.24 | –0.51 | .610 | .776 |
| | MCI Aβ+ – SCD Aβ+ | –0.45 [–1.05; 0.15] | 0.23 | –1.99 | .048* | .084 |
| | DAT Aβ+ – MCI Aβ+ | –1.07 [–1.87; –0.27] | 0.30 | –3.52 | < .001*** | .002** |

Abbreviations: BA, Brodmann area. CA, cornu ammonis. CA23DG, cornu ammonis 2, 3, and dentate gyrus. DAT, dementia of the Alzheimer type. HC, healthy control. MCI, mild cognitive impairment. SCD, subjective cognitive decline.

**Supplementary Table 4** Summary statistics of the fixed effects in linear mixed effects models predicting structural MRI markers, derived from linear mixed effects models.

| Marker | $R^2_{conditional}$ | $R^2_{marginal}$ | Predictor | *b* [95% C.I.] | *SE* | *t* | *df* | $\eta^2_{partial}$ | *p* |
|---|---|---|---|---|---|---|---|---|---|
| Amygdala, volume | 0.99 | 0.46 | Intercept | 3.97 [2.32; 5.63] | 0.85 | 4.70 | 331.03 | 0.06 | < .001*** |
| | | | Age, years | –0.06 [–0.08; –0.03] | 0.01 | –4.75 | 331.01 | 0.06 | < .001*** |
| | | | Years since baseline | 0.12 [–0.05; 0.29] | 0.09 | 1.43 | 266.74 | 0.01 | .153 |
| | | | Sex, male | 0.12 [–0.15; 0.39] | 0.14 | 0.89 | 330.92 | 0.00 | .372 |
| | | | HC Aβ+ diagnosis | 0.05 [–0.47; 0.56] | 0.26 | 0.18 | 330.43 | 0.00 | .857 |
| | | | SCD Aβ+ diagnosis | –0.71 [–1.07; –0.34] | 0.19 | –3.81 | 330.93 | 0.04 | < .001*** |
| | | | MCI Aβ+ diagnosis | –1.60 [–2.01; –1.18] | 0.21 | –7.55 | 331.02 | 0.15 | < .001*** |
| | | | DAT Aβ+ diagnosis | –2.99 [–3.47; –2.52] | 0.24 | –12.42 | 331.55 | 0.32 | < .001*** |
| | | | Age, years × years since baseline | 0.00 [0.00; 0.00] | 0.00 | –1.87 | 266.73 | 0.01 | .063 |
| | | | Sex, male × years since baseline | –0.01 [–0.04; 0.02] | 0.01 | –0.62 | 257.62 | 0.00 | .538 |
| | | | HC Aβ+ diagnosis × years since baseline | –0.07 [–0.12; –0.03] | 0.02 | –2.99 | 224.83 | 0.04 | .003** |
| | | | SCD Aβ+ diagnosis × years since baseline | –0.05 [–0.09; –0.01] | 0.02 | –2.70 | 254.74 | 0.03 | .007** |
| | | | MCI Aβ+ diagnosis × years since baseline | –0.11 [–0.16; –0.07] | 0.02 | –5.19 | 272.70 | 0.09 | < .001*** |
| | | | DAT Aβ+ diagnosis × years since baseline | –0.14 [–0.19; –0.08] | 0.03 | –4.90 | 347.63 | 0.06 | < .001*** |
| CA1, volume | 0.98 | 0.27 | Intercept | 2.75 [1.23; 4.27] | 0.78 | 3.55 | 283.05 | 0.04 | < .001*** |
| | | | Age, years | –0.04 [–0.06; –0.02] | 0.01 | –3.51 | 283.06 | 0.04 | < .001*** |
| | | | Years since baseline | 0.13 [–0.02; 0.28] | 0.08 | 1.64 | 257.78 | 0.01 | .102 |
| | | | Sex, male | –0.06 [–0.32; 0.19] | 0.13 | –0.50 | 282.87 | 0.00 | .620 |
| | | | HC Aβ+ diagnosis | 0.35 [–0.08; 0.79] | 0.22 | 1.60 | 282.61 | 0.01 | .110 |
| | | | SCD Aβ+ diagnosis | –0.37 [–0.71; –0.04] | 0.17 | –2.17 | 282.95 | 0.02 | .031* |
| | | | MCI Aβ+ diagnosis | –0.83 [–1.25; –0.42] | 0.21 | –3.94 | 283.17 | 0.05 | < .001*** |
| | | | DAT Aβ+ diagnosis | –1.88 [–2.40; –1.35] | 0.27 | –7.01 | 283.47 | 0.15 | < .001*** |
| | | | Age, years × years since baseline | 0.00 [0.00; 0.00] | 0.00 | –1.83 | 257.06 | 0.01 | .069 |
| | | | Sex, male × years since baseline | 0.00 [–0.03; 0.02] | 0.01 | –0.27 | 246.36 | 0.00 | .787 |
| | | | HC Aβ+ diagnosis × years since baseline | –0.02 [–0.06; 0.02] | 0.02 | –1.11 | 229.85 | 0.01 | .269 |
| | | | SCD Aβ+ diagnosis × years since baseline | –0.02 [–0.05; 0.02] | 0.02 | –0.99 | 252.49 | 0.00 | .325 |
| | | | MCI Aβ+ diagnosis × years since baseline | –0.05 [–0.09; 0.00] | 0.02 | –2.13 | 280.11 | 0.02 | .034* |
| | | | DAT Aβ+ diagnosis × years since baseline | –0.08 [–0.13; –0.02] | 0.03 | –2.63 | 319.87 | 0.02 | .009** |
| CA23DG, volume | 0.98 | 0.18 | Intercept | 1.89 [0.47; 3.32] | 0.73 | 2.60 | 282.85 | 0.02 | .010** |
| | | | Age, years | –0.03 [–0.05; –0.01] | 0.01 | –2.53 | 282.85 | 0.02 | .012* |
| | | | Years since baseline | 0.10 [–0.05; 0.25] | 0.08 | 1.33 | 251.68 | 0.01 | .185 |
| | | | Sex, male | –0.15 [–0.38; 0.09] | 0.12 | –1.23 | 282.67 | 0.01 | .221 |
| | | | HC Aβ+ diagnosis | –0.02 [–0.43; 0.39] | 0.21 | –0.09 | 282.38 | 0.00 | .926 |
| | | | SCD Aβ+ diagnosis | –0.40 [–0.71; –0.08] | 0.16 | –2.47 | 282.74 | 0.02 | .014* |
| | | | MCI Aβ+ diagnosis | –0.59 [–0.98; –0.20] | 0.20 | –2.97 | 282.98 | 0.03 | .003** |
| | | | DAT Aβ+ diagnosis | –1.21 [–1.70; –0.71] | 0.25 | –4.81 | 283.30 | 0.08 | < .001*** |
| | | | Age, years × years since baseline | 0.00 [0.00; 0.00] | 0.00 | –2.19 | 250.99 | 0.02 | .029* |
| | | | Sex, male × years since baseline | 0.00 [–0.02; 0.02] | 0.01 | –0.07 | 240.30 | 0.00 | .946 |
| | | | HC Aβ+ diagnosis × years since baseline | –0.01 [–0.04; 0.03] | 0.02 | –0.28 | 223.93 | 0.00 | .783 |
| | | | SCD Aβ+ diagnosis × years since baseline | –0.03 [–0.06; 0.00] | 0.02 | –1.86 | 246.51 | 0.01 | .064 |
| | | | MCI Aβ+ diagnosis × years since baseline | –0.05 [–0.09; –0.01] | 0.02 | –2.30 | 273.51 | 0.02 | .022* |
| | | | DAT Aβ+ diagnosis × years since baseline | –0.10 [–0.15; –0.04] | 0.03 | –3.50 | 313.44 | 0.04 | < .001*** |
| Subiculum, volume | 0.99 | 0.37 | Intercept | 4.39 [2.95; 5.83] | 0.74 | 5.96 | 283.27 | 0.11 | < .001*** |
| | | | Age, years | –0.07 [–0.09; –0.04] | 0.01 | –6.04 | 283.27 | 0.11 | < .001*** |
| | | | Years since baseline | 0.17 [0.02; 0.32] | 0.08 | 2.19 | 252.45 | 0.02 | .029* |
| | | | Sex, male | 0.19 [–0.05; 0.43] | 0.12 | 1.58 | 283.09 | 0.01 | .114 |
| | | | HC Aβ+ diagnosis | 0.16 [–0.25; 0.58] | 0.21 | 0.78 | 282.82 | 0.00 | .435 |
| | | | SCD Aβ+ diagnosis | –0.44 [–0.76; –0.12] | 0.16 | –2.70 | 283.17 | 0.03 | .007** |
| | | | MCI Aβ+ diagnosis | –0.91 [–1.31; –0.52] | 0.20 | –4.55 | 283.39 | 0.07 | < .001*** |
| | | | DAT Aβ+ diagnosis | –2.04 [–2.54; –1.54] | 0.25 | –8.02 | 283.70 | 0.18 | < .001*** |
| | | | Age, years × years since baseline | 0.00 [–0.01; 0.00] | 0.00 | –2.67 | 251.85 | 0.03 | .008** |
| | | | Sex, male × years since baseline | –0.01 [–0.03; 0.02] | 0.01 | –0.74 | 241.00 | 0.00 | .459 |
| | | | HC Aβ+ diagnosis × years since baseline | –0.02 [–0.06; 0.03] | 0.02 | –0.74 | 224.70 | 0.00 | .463 |
| | | | SCD Aβ+ diagnosis × years since baseline | –0.05 [–0.08; –0.02] | 0.02 | –2.91 | 247.55 | 0.03 | .004** |
| | | | MCI Aβ+ diagnosis × years since baseline | –0.07 [–0.11; –0.03] | 0.02 | –3.13 | 273.29 | 0.03 | .002** |
| | | | DAT Aβ+ diagnosis × years since baseline | –0.12 [–0.17; –0.06] | 0.03 | –3.93 | 313.12 | 0.05 | < .001*** |
| | 0.97 | 0.35 | Intercept | 4.49 [3.08; 5.91] | 0.72 | 6.22 | 283.33 | 0.12 | < .001*** |
| | | | Age, years | –0.07 [–0.09; –0.04] | 0.01 | –6.21 | 283.35 | 0.12 | < .001*** |

| | | | | | | | | | |
|---|---|---|---|---|---|---|---|---|---|
| Hippocampal tail, volume | | | Years since baseline | 0.14 [–0.06; 0.34] | 0.10 | 1.39 | 247.63 | 0.01 | .165 |
| | | | Sex, male | –0.04 [–0.27; 0.19] | 0.12 | –0.34 | 282.99 | 0.00 | .738 |
| | | | HC Aβ+ diagnosis | –0.09 [–0.49; 0.31] | 0.21 | –0.44 | 282.48 | 0.00 | .660 |
| | | | SCD Aβ+ diagnosis | –0.35 [–0.67; –0.04] | 0.16 | –2.22 | 283.14 | 0.02 | .027* |
| | | | MCI Aβ+ diagnosis | –0.95 [–1.34; –0.57] | 0.20 | –4.83 | 283.56 | 0.08 | < .001*** |
| | | | DAT Aβ+ diagnosis | –1.77 [–2.26; –1.28] | 0.25 | –7.11 | 284.16 | 0.15 | < .001*** |
| | | | Age, years × years since baseline | 0.00 [–0.01; 0.00] | 0.00 | –1.57 | 246.97 | 0.01 | .119 |
| | | | Sex, male × years since baseline | –0.02 [–0.05; 0.01] | 0.02 | –1.05 | 236.29 | 0.00 | .293 |
| | | | HC Aβ+ diagnosis × years since baseline | –0.01 [–0.07; 0.04] | 0.03 | –0.50 | 220.04 | 0.00 | .614 |
| | | | SCD Aβ+ diagnosis × years since baseline | –0.05 [–0.09; 0.00] | 0.02 | –2.12 | 242.57 | 0.02 | .035* |
| | | | MCI Aβ+ diagnosis × years since baseline | –0.03 [–0.09; 0.02] | 0.03 | –1.12 | 269.10 | 0.00 | .263 |
| | | | DAT Aβ+ diagnosis × years since baseline | –0.04 [–0.12; 0.03] | 0.04 | –1.09 | 308.93 | 0.00 | .276 |
| Entorhinal cortex, volume | 0.97 | 0.24 | Intercept | 2.23 [0.81; 3.66] | 0.72 | 3.08 | 283.23 | 0.03 | .002** |
| | | | Age, years | –0.03 [–0.05; –0.01] | 0.01 | –3.10 | 283.24 | 0.03 | .002** |
| | | | Years since baseline | 0.25 [0.08; 0.43] | 0.09 | 2.91 | 253.61 | 0.03 | .004** |
| | | | Sex, male | 0.08 [–0.16; 0.31] | 0.12 | 0.65 | 282.98 | 0.00 | .515 |
| | | | HC Aβ+ diagnosis | –0.03 [–0.44; 0.37] | 0.21 | –0.17 | 282.60 | 0.00 | .866 |
| | | | SCD Aβ+ diagnosis | –0.47 [–0.78; –0.15] | 0.16 | –2.92 | 283.09 | 0.03 | .004** |
| | | | MCI Aβ+ diagnosis | –0.77 [–1.16; –0.39] | 0.20 | –3.91 | 283.40 | 0.05 | < .001*** |
| | | | DAT Aβ+ diagnosis | –1.42 [–1.91; –0.92] | 0.25 | –5.65 | 283.83 | 0.10 | < .001*** |
| | | | Age, years × years since baseline | 0.00 [–0.01; 0.00] | 0.00 | –2.94 | 252.92 | 0.03 | .004** |
| | | | Sex, male × years since baseline | –0.01 [–0.03; 0.02] | 0.01 | –0.45 | 242.20 | 0.00 | .653 |
| | | | HC Aβ+ diagnosis × years since baseline | –0.07 [–0.12; –0.02] | 0.02 | –3.01 | 225.81 | 0.04 | .003** |
| | | | SCD Aβ+ diagnosis × years since baseline | –0.05 [–0.08; –0.01] | 0.02 | –2.42 | 248.46 | 0.02 | .016* |
| | | | MCI Aβ+ diagnosis × years since baseline | –0.08 [–0.13; –0.03] | 0.02 | –3.30 | 275.28 | 0.04 | .001** |
| | | | DAT Aβ+ diagnosis × years since baseline | –0.17 [–0.24; –0.11] | 0.03 | –5.24 | 315.30 | 0.08 | < .001*** |
| BA35, volume | 0.97 | 0.13 | Intercept | 2.43 [1.01; 3.86] | 0.73 | 3.35 | 283.14 | 0.04 | < .001*** |
| | | | Age, years | –0.04 [–0.06; –0.02] | 0.01 | –3.41 | 283.15 | 0.04 | < .001*** |
| | | | Years since baseline | 0.19 [0.01; 0.37] | 0.09 | 2.02 | 258.01 | 0.02 | .044* |
| | | | Sex, male | 0.15 [–0.09; 0.38] | 0.12 | 1.22 | 282.88 | 0.01 | .224 |
| | | | HC Aβ+ diagnosis | 0.15 [–0.26; 0.55] | 0.21 | 0.71 | 282.50 | 0.00 | .478 |
| | | | SCD Aβ+ diagnosis | 0.05 [–0.26; 0.37] | 0.16 | 0.33 | 283.00 | 0.00 | .745 |
| | | | MCI Aβ+ diagnosis | –0.22 [–0.61; 0.17] | 0.20 | –1.11 | 283.32 | 0.00 | .268 |
| | | | DAT Aβ+ diagnosis | –0.82 [–1.31; –0.33] | 0.25 | –3.28 | 283.76 | 0.04 | .001** |
| | | | Age, years × years since baseline | 0.00 [–0.01; 0.00] | 0.00 | –2.29 | 257.43 | 0.02 | .023* |
| | | | Sex, male × years since baseline | –0.01 [–0.04; 0.02] | 0.01 | –0.39 | 246.40 | 0.00 | .695 |
| | | | HC Aβ+ diagnosis × years since baseline | –0.07 [–0.12; –0.02] | 0.02 | –2.73 | 229.91 | 0.03 | .007** |
| | | | SCD Aβ+ diagnosis × years since baseline | –0.04 [–0.08; 0.00] | 0.02 | –2.04 | 253.17 | 0.02 | .043* |
| | | | MCI Aβ+ diagnosis × years since baseline | –0.04 [–0.09; 0.01] | 0.03 | –1.69 | 278.95 | 0.01 | .093 |
| | | | DAT Aβ+ diagnosis × years since baseline | –0.16 [–0.22; –0.09] | 0.03 | –4.49 | 318.92 | 0.06 | < .001*** |
| BA36, volume | 0.99 | 0.07 | Intercept | 1.81 [0.21; 3.42] | 0.82 | 2.21 | 283.03 | 0.02 | .028* |
| | | | Age, years | –0.03 [–0.05; 0.00] | 0.01 | –2.28 | 283.03 | 0.02 | .023* |
| | | | Years since baseline | 0.07 [–0.05; 0.20] | 0.06 | 1.13 | 223.34 | 0.01 | .259 |
| | | | Sex, male | 0.17 [–0.09; 0.44] | 0.14 | 1.27 | 282.93 | 0.01 | .204 |
| | | | HC Aβ+ diagnosis | 0.17 [–0.29; 0.63] | 0.23 | 0.72 | 282.77 | 0.00 | .473 |
| | | | SCD Aβ+ diagnosis | 0.05 [–0.31; 0.40] | 0.18 | 0.25 | 282.97 | 0.00 | .799 |
| | | | MCI Aβ+ diagnosis | –0.14 [–0.58; 0.29] | 0.22 | –0.65 | 283.10 | 0.00 | .519 |
| | | | DAT Aβ+ diagnosis | –0.62 [–1.18; –0.07] | 0.28 | –2.19 | 283.28 | 0.02 | .029* |
| | | | Age, years × years since baseline | 0.00 [0.00; 0.00] | 0.00 | –1.47 | 222.70 | 0.01 | .142 |
| | | | Sex, male × years since baseline | 0.00 [–0.02; 0.02] | 0.01 | –0.04 | 212.59 | 0.00 | .968 |
| | | | HC Aβ+ diagnosis × years since baseline | –0.03 [–0.06; 0.00] | 0.02 | –1.76 | 197.24 | 0.02 | .080 |
| | | | SCD Aβ+ diagnosis × years since baseline | –0.03 [–0.06; –0.01] | 0.01 | –2.37 | 218.48 | 0.03 | .019* |
| | | | MCI Aβ+ diagnosis × years since baseline | –0.04 [–0.08; –0.01] | 0.02 | –2.50 | 244.15 | 0.03 | .013* |
| | | | DAT Aβ+ diagnosis × years since baseline | –0.18 [–0.23; –0.13] | 0.02 | –7.51 | 282.73 | 0.17 | < .001*** |
| Parahippocampal cortex, volume[1] | 0.97 | 0.07 | Intercept | 2.22 [0.57; 3.87] | 0.84 | 2.63 | 283.35 | 0.02 | .009** |
| | | | Age, years | –0.03 [–0.06; –0.01] | 0.01 | –2.68 | 283.36 | 0.02 | .008** |
| | | | Years since baseline | 0.05 [–0.12; 0.22] | 0.09 | 0.61 | 217.77 | 0.00 | .543 |
| | | | Sex, male | 0.15 [–0.13; 0.42] | 0.14 | 1.06 | 283.13 | 0.00 | .289 |
| | | | HC Aβ+ diagnosis | 0.26 [–0.22; 0.73] | 0.24 | 1.06 | 282.79 | 0.00 | .288 |
| | | | SCD Aβ+ diagnosis | –0.01 [–0.37; 0.36] | 0.19 | –0.05 | 283.22 | 0.00 | .961 |
| | | | MCI Aβ+ diagnosis | –0.29 [–0.74; 0.16] | 0.23 | –1.26 | 283.50 | 0.01 | .208 |

| | | | | | | | | | |
|---|---|---|---|---|---|---|---|---|---|
| | | | DAT Aβ+ diagnosis | −0.34 [−0.91; 0.23] | 0.29 | −1.16 | 283.88 | 0.00 | .246 |
| | | | Age, years × years since baseline | 0.00 [0.00; 0.00] | 0.00 | −1.16 | 217.15 | 0.01 | .249 |
| | | | Sex, male × years since baseline | −0.02 [−0.05; 0.01] | 0.01 | −1.44 | 207.41 | 0.01 | .151 |
| | | | HC Aβ+ diagnosis × years since baseline | 0.03 [−0.02; 0.07] | 0.02 | 1.17 | 192.71 | 0.01 | .243 |
| | | | SCD Aβ+ diagnosis × years since baseline | −0.01 [−0.05; 0.03] | 0.02 | −0.49 | 213.02 | 0.00 | .624 |
| | | | MCI Aβ+ diagnosis × years since baseline | −0.03 [−0.08; 0.01] | 0.02 | −1.44 | 240.06 | 0.01 | .151 |
| | | | DAT Aβ+ diagnosis × years since baseline | −0.14 [−0.21; −0.08] | 0.03 | −4.32 | 274.19 | 0.06 | < .001*** |
| Retrosplenial cortex, average thickness | 0.96 | 0.14 | Intercept | 2.39 [0.98; 3.81] | 0.72 | 3.31 | 283.23 | 0.04 | .001** |
| | | | Age, years | −0.04 [−0.06; −0.01] | 0.01 | −3.32 | 283.23 | 0.04 | .001** |
| | | | Years since baseline | 0.03 [−0.14; 0.20] | 0.09 | 0.34 | 251.39 | 0.00 | .734 |
| | | | Sex, male | −0.01 [−0.24; 0.23] | 0.12 | −0.04 | 282.99 | 0.00 | .965 |
| | | | HC Aβ+ diagnosis | −0.14 [−0.55; 0.26] | 0.21 | −0.70 | 282.54 | 0.00 | .483 |
| | | | SCD Aβ+ diagnosis | −0.08 [−0.40; 0.23] | 0.16 | −0.53 | 283.22 | 0.00 | .598 |
| | | | MCI Aβ+ diagnosis | −0.41 [−0.80; −0.02] | 0.20 | −2.08 | 283.30 | 0.02 | .038* |
| | | | DAT Aβ+ diagnosis | −0.90 [−1.39; −0.41] | 0.25 | −3.59 | 283.93 | 0.04 | < .001*** |
| | | | Age, years × years since baseline | 0.00 [0.00; 0.00] | 0.00 | −0.72 | 250.62 | 0.00 | .469 |
| | | | Sex, male × years since baseline | 0.00 [−0.03; 0.03] | 0.01 | 0.03 | 239.93 | 0.00 | .973 |
| | | | HC Aβ+ diagnosis × years since baseline | 0.01 [−0.04; 0.05] | 0.02 | 0.25 | 221.92 | 0.00 | .804 |
| | | | SCD Aβ+ diagnosis × years since baseline | −0.04 [−0.07; 0.00] | 0.02 | −1.87 | 249.67 | 0.01 | .063 |
| | | | MCI Aβ+ diagnosis × years since baseline | −0.06 [−0.11; −0.02] | 0.02 | −2.67 | 257.60 | 0.03 | .008** |
| | | | DAT Aβ+ diagnosis × years since baseline | −0.16 [−0.22; −0.09] | 0.03 | −4.90 | 282.91 | 0.08 | < .001*** |
| Posterior cingulate cortex, average thickness | 0.89 | 0.08 | Intercept | 1.52 [0.04; 3.00] | 0.75 | 2.01 | 316.86 | 0.01 | .045* |
| | | | Age, years | −0.02 [−0.04; 0.00] | 0.01 | −1.96 | 316.85 | 0.01 | .051 |
| | | | Years since baseline | 0.20 [−0.07; 0.47] | 0.14 | 1.45 | 721.93 | 0.00 | .147 |
| | | | Sex, male | −0.13 [−0.37; 0.12] | 0.12 | −1.02 | 316.17 | 0.00 | .308 |
| | | | HC Aβ+ diagnosis | 0.01 [−0.41; 0.43] | 0.21 | 0.04 | 314.83 | 0.00 | .971 |
| | | | SCD Aβ+ diagnosis | −0.11 [−0.44; 0.22] | 0.17 | −0.66 | 316.83 | 0.00 | .507 |
| | | | MCI Aβ+ diagnosis | −0.28 [−0.68; 0.13] | 0.21 | −1.34 | 317.07 | 0.01 | .180 |
| | | | DAT Aβ+ diagnosis | −0.51 [−1.02; 0.00] | 0.26 | −1.96 | 318.89 | 0.01 | .051 |
| | | | Age, years × years since baseline | 0.00 [−0.01; 0.00] | 0.00 | −1.38 | 722.19 | 0.00 | .169 |
| | | | Sex, male × years since baseline | −0.02 [−0.06; 0.02] | 0.02 | −1.01 | 720.27 | 0.00 | .315 |
| | | | HC Aβ+ diagnosis × years since baseline | 0.02 [−0.05; 0.09] | 0.04 | 0.52 | 718.77 | 0.00 | .601 |
| | | | SCD Aβ+ diagnosis × years since baseline | −0.02 [−0.08; 0.04] | 0.03 | −0.69 | 721.56 | 0.00 | .490 |
| | | | MCI Aβ+ diagnosis × years since baseline | −0.05 [−0.13; 0.02] | 0.04 | −1.40 | 721.81 | 0.00 | .162 |
| | | | DAT Aβ+ diagnosis × years since baseline | −0.25 [−0.35; −0.15] | 0.05 | −4.92 | 724.29 | 0.03 | < .001*** |
| Precuneus, average thickness | 0.94 | 0.25 | Intercept | 2.67 [1.22; 4.12] | 0.74 | 3.62 | 283.22 | 0.04 | < .001*** |
| | | | Age, years | −0.04 [−0.06; −0.02] | 0.01 | −3.54 | 283.21 | 0.04 | < .001*** |
| | | | Years since baseline | −0.04 [−0.33; 0.26] | 0.15 | −0.24 | 265.24 | 0.00 | .814 |
| | | | Sex, male | −0.12 [−0.36; 0.12] | 0.12 | −1.01 | 282.65 | 0.00 | .311 |
| | | | HC Aβ+ diagnosis | 0.40 [−0.01; 0.81] | 0.21 | 1.92 | 281.63 | 0.01 | .056 |
| | | | SCD Aβ+ diagnosis | −0.09 [−0.41; 0.22] | 0.16 | −0.58 | 283.18 | 0.00 | .564 |
| | | | MCI Aβ+ diagnosis | −0.62 [−1.02; −0.23] | 0.20 | −3.10 | 283.37 | 0.03 | .002** |
| | | | DAT Aβ+ diagnosis | −1.38 [−1.88; −0.88] | 0.26 | −5.39 | 284.78 | 0.09 | < .001*** |
| | | | Age, years × years since baseline | 0.00 [0.00; 0.00] | 0.00 | −0.09 | 264.58 | 0.00 | .925 |
| | | | Sex, male × years since baseline | −0.04 [−0.09; 0.01] | 0.02 | −1.69 | 252.59 | 0.01 | .092 |
| | | | HC Aβ+ diagnosis × years since baseline | −0.02 [−0.10; 0.06] | 0.04 | −0.48 | 234.62 | 0.00 | .631 |
| | | | SCD Aβ+ diagnosis × years since baseline | −0.10 [−0.17; −0.04] | 0.03 | −3.10 | 262.36 | 0.04 | .002** |
| | | | MCI Aβ+ diagnosis × years since baseline | −0.11 [−0.20; −0.03] | 0.04 | −2.78 | 270.59 | 0.03 | .006** |
| | | | DAT Aβ+ diagnosis × years since baseline | −0.32 [−0.43; −0.22] | 0.06 | −5.89 | 298.62 | 0.10 | < .001*** |
| Infertior parietal cortex, average thickness | 0.93 | 0.30 | Intercept | 3.86 [2.35; 5.36] | 0.77 | 5.03 | 283.37 | 0.08 | < .001*** |
| | | | Age, years | −0.05 [−0.08; −0.03] | 0.01 | −4.87 | 283.36 | 0.08 | < .001*** |
| | | | Years since baseline | −0.03 [−0.38; 0.32] | 0.18 | −0.16 | 275.11 | 0.00 | .870 |
| | | | Sex, male | −0.28 [−0.53; −0.03] | 0.13 | −2.19 | 282.65 | 0.02 | .029* |
| | | | HC Aβ+ diagnosis | 0.07 [−0.36; 0.50] | 0.22 | 0.32 | 281.39 | 0.00 | .749 |
| | | | SCD Aβ+ diagnosis | −0.06 [−0.39; 0.27] | 0.17 | −0.36 | 283.31 | 0.00 | .716 |
| | | | MCI Aβ+ diagnosis | −0.52 [−0.93; −0.11] | 0.21 | −2.47 | 283.55 | 0.02 | .014* |
| | | | DAT Aβ+ diagnosis | −1.51 [−2.03; −0.99] | 0.27 | −5.68 | 285.32 | 0.10 | < .001*** |
| | | | Age, years × years since baseline | 0.00 [−0.01; 0.00] | 0.00 | −0.20 | 274.50 | 0.00 | .844 |
| | | | Sex, male × years since baseline | −0.03 [−0.08; 0.03] | 0.03 | −0.90 | 262.07 | 0.00 | .367 |

| | | | | | | | | | |
|---|---|---|---|---|---|---|---|---|---|
| | | | HC Aβ+ diagnosis × years since baseline | 0.00 [–0.10; 0.09] | 0.05 | –0.10 | 243.96 | 0.00 | .919 |
| | | | SCD Aβ+ diagnosis × years since baseline | –0.15 [–0.23; –0.08] | 0.04 | –3.90 | 272.01 | 0.05 | < .001*** |
| | | | MCI Aβ+ diagnosis × years since baseline | –0.16 [–0.25; –0.06] | 0.05 | –3.17 | 280.32 | 0.03 | .002** |
| | | | DAT Aβ+ diagnosis × years since baseline | –0.52 [–0.65; –0.39] | 0.07 | –8.00 | 308.72 | 0.17 | < .001*** |

[1]Fitted without random slopes due to insufficient variance in random slope estimates.

Abbreviations: AMY, amygdala. BA, Brodmann area. CA, cornu ammonis. CA23DG, cornu ammonis 2, 3, and dentate gyrus. DAT, dementia of the Alzheimer type. ERC, entorhinal cortex. HC, healthy control. IPC, inferior parietal cortex. MCI, mild cognitive impairment. PCC, posterior cingulate cortex. PHC, parahippocampal cortex. PRE, precuneus. RSC, retrosplenial cortex. SCD, subjective cognitive decline. SUB, subiculum. TAIL, hippocampal tail.

**Supplementary Table 5** Estimated marginal means of the linear fixed effect of years since baseline on structural MRI markers for each diagnostic group, derived from linear mixed effects models.

| Marker | Diagnostic group | *b* [95% C.I.] | *SE* | *t* | *df* | $\eta^2_{partial}$ | *p* |
|---|---|---|---|---|---|---|---|
| Amygdala, volume | HC Aβ– | –0.05 [–0.07; –0.03] | 0.01 | –4.74 | 257.94 | 0.08 | < .001*** |
| | HC Aβ + | –0.12 [–0.17; –0.08] | 0.02 | –5.46 | 247.84 | 0.11 | < .001*** |
| | SCD Aβ + | –0.10 [–0.13; –0.07] | 0.01 | –6.58 | 294.48 | 0.13 | < .001*** |
| | MCI Aβ + | –0.16 [–0.20; –0.12] | 0.02 | –8.67 | 310.15 | 0.20 | < .001*** |
| | DAT Aβ + | –0.18 [–0.23; –0.14] | 0.03 | –7.30 | 408.62 | 0.12 | < .001*** |
| CA1, volume | HC Aβ– | –0.02 [–0.04; 0.00] | 0.01 | –2.45 | 223.69 | 0.03 | .015* |
| | HC Aβ + | –0.04 [–0.08; –0.01] | 0.02 | –2.32 | 224.92 | 0.02 | .021* |
| | SCD Aβ + | –0.04 [–0.06; –0.01] | 0.01 | –2.58 | 258.92 | 0.03 | .010* |
| | MCI Aβ + | –0.07 [–0.10; –0.03] | 0.02 | –3.36 | 283.99 | 0.04 | < .001*** |
| | DAT Aβ + | –0.10 [–0.15; –0.04] | 0.03 | –3.49 | 324.38 | 0.04 | < .001*** |
| CA23DG, volume | HC Aβ– | –0.07 [–0.09; –0.05] | 0.01 | –8.67 | 224.76 | 0.25 | < .001*** |
| | HC Aβ + | –0.08 [–0.11; –0.04] | 0.02 | –4.17 | 226.03 | 0.07 | < .001*** |
| | SCD Aβ + | –0.10 [–0.13; –0.07] | 0.01 | –7.22 | 260.35 | 0.17 | < .001*** |
| | MCI Aβ + | –0.12 [–0.16; –0.08] | 0.02 | –6.17 | 284.96 | 0.12 | < .001*** |
| | DAT Aβ + | –0.17 [–0.22; –0.12] | 0.03 | –6.27 | 325.70 | 0.11 | < .001*** |
| Subiculum, volume | HC Aβ– | –0.05 [–0.06; –0.03] | 0.01 | –5.71 | 227.59 | 0.13 | < .001*** |
| | HC Aβ + | –0.06 [–0.10; –0.03] | 0.02 | –3.36 | 229.01 | 0.05 | < .001*** |
| | SCD Aβ + | –0.10 [–0.13; –0.07] | 0.01 | –6.77 | 263.99 | 0.15 | < .001*** |
| | MCI Aβ + | –0.12 [–0.16; –0.08] | 0.02 | –5.86 | 287.17 | 0.11 | < .001*** |
| | DAT Aβ + | –0.16 [–0.22; –0.11] | 0.03 | –5.86 | 328.03 | 0.09 | < .001*** |
| Hippocampal tail, volume | HC Aβ– | –0.03 [–0.05; –0.01] | 0.01 | –2.83 | 225.33 | 0.03 | .005** |
| | HC Aβ + | –0.04 [–0.09; 0.00] | 0.02 | –1.82 | 226.62 | 0.01 | .070 |
| | SCD Aβ + | –0.08 [–0.12; –0.04] | 0.02 | –4.15 | 261.11 | 0.06 | < .001*** |
| | MCI Aβ + | –0.06 [–0.11; –0.01] | 0.03 | –2.42 | 285.47 | 0.02 | .016* |
| | DAT Aβ + | –0.07 [–0.14; 0.00] | 0.04 | –2.00 | 326.34 | 0.01 | .047* |
| Entorhinal cortex, volume | HC Aβ– | –0.01 [–0.03; 0.00] | 0.01 | –1.44 | 225.36 | 0.01 | .150 |
| | HC Aβ + | –0.08 [–0.13; –0.04] | 0.02 | –3.97 | 226.65 | 0.06 | < .001*** |
| | SCD Aβ + | –0.06 [–0.09; –0.03] | 0.02 | –3.70 | 261.14 | 0.05 | < .001*** |
| | MCI Aβ + | –0.09 [–0.14; –0.05] | 0.02 | –4.22 | 285.48 | 0.06 | < .001*** |
| | DAT Aβ + | –0.19 [–0.25; –0.12] | 0.03 | –5.94 | 326.35 | 0.10 | < .001*** |
| BA35, volume | HC Aβ– | –0.03 [–0.05; –0.01] | 0.01 | –3.36 | 228.50 | 0.05 | < .001*** |
| | HC Aβ + | –0.10 [–0.15; –0.06] | 0.02 | –4.51 | 230.08 | 0.08 | < .001*** |
| | SCD Aβ + | –0.08 [–0.11; –0.04] | 0.02 | –4.37 | 265.53 | 0.07 | < .001*** |
| | MCI Aβ + | –0.08 [–0.12; –0.03] | 0.02 | –3.28 | 288.40 | 0.04 | .001** |
| | DAT Aβ + | –0.19 [–0.25; –0.13] | 0.03 | –5.74 | 329.24 | 0.09 | < .001*** |
| BA36, volume | HC Aβ– | –0.02 [–0.04; –0.01] | 0.01 | –3.60 | 224.66 | 0.05 | < .001*** |
| | HC Aβ + | –0.05 [–0.08; –0.02] | 0.02 | –3.55 | 225.93 | 0.05 | < .001*** |
| | SCD Aβ + | –0.06 [–0.08; –0.03] | 0.01 | –4.89 | 260.22 | 0.08 | < .001*** |
| | MCI Aβ + | –0.07 [–0.10; –0.04] | 0.02 | –4.26 | 284.88 | 0.06 | < .001*** |
| | DAT Aβ + | –0.20 [–0.25; –0.16] | 0.02 | –8.97 | 325.56 | 0.20 | < .001*** |
| Parahippocampal cortex, volume[1] | HC Aβ– | –0.06 [–0.08; –0.04] | 0.01 | –6.51 | 218.75 | 0.16 | < .001*** |
| | HC Aβ + | –0.03 [–0.07; 0.01] | 0.02 | –1.61 | 220.15 | 0.01 | .109 |
| | SCD Aβ + | –0.07 [–0.10; –0.04] | 0.02 | –4.31 | 252.56 | 0.07 | < .001*** |
| | MCI Aβ + | –0.09 [–0.14; –0.05] | 0.02 | –4.27 | 279.03 | 0.06 | < .001*** |
| | DAT Aβ + | –0.20 [–0.26; –0.14] | 0.03 | –6.42 | 314.76 | 0.12 | < .001*** |
| Retrosplenial cortex, average thickness | HC Aβ– | –0.04 [–0.05; –0.02] | 0.01 | –3.76 | 228.92 | 0.06 | < .001*** |
| | HC Aβ + | –0.03 [–0.07; 0.01] | 0.02 | –1.42 | 223.58 | 0.01 | .157 |
| | SCD Aβ + | –0.07 [–0.10; –0.04] | 0.02 | –4.42 | 262.97 | 0.07 | < .001*** |
| | MCI Aβ + | –0.10 [–0.14; –0.06] | 0.02 | –4.58 | 264.02 | 0.07 | < .001*** |
| | DAT Aβ + | –0.19 [–0.25; –0.13] | 0.03 | –6.32 | 291.34 | 0.12 | < .001*** |
| Posterior cingulate cortex, average thickness | HC Aβ– | –0.01 [–0.04; 0.02] | 0.01 | –0.46 | 717.73 | 0.00 | .647 |
| | HC Aβ + | 0.01 [–0.05; 0.08] | 0.03 | 0.37 | 718.61 | 0.00 | .710 |
| | SCD Aβ + | –0.03 [–0.08; 0.02] | 0.03 | –1.09 | 722.76 | 0.00 | .278 |
| | MCI Aβ + | –0.06 [–0.13; 0.01] | 0.03 | –1.74 | 721.88 | 0.00 | .081 |
| | DAT Aβ + | –0.26 [–0.35; –0.16] | 0.05 | –5.32 | 724.34 | 0.04 | < .001*** |
| Precuneus, average thickness | HC Aβ– | –0.07 [–0.10; –0.04] | 0.02 | –4.32 | 235.45 | 0.07 | < .001*** |
| | HC Aβ + | –0.09 [–0.16; –0.02] | 0.04 | –2.47 | 230.97 | 0.03 | .014* |
| | SCD Aβ + | –0.17 [–0.23; –0.12] | 0.03 | –6.25 | 270.64 | 0.13 | < .001*** |
| | MCI Aβ + | –0.19 [–0.26; –0.11] | 0.04 | –4.97 | 271.80 | 0.08 | < .001*** |
| | DAT Aβ + | –0.40 [–0.50; –0.29] | 0.05 | –7.57 | 301.82 | 0.16 | < .001*** |
| Inferior parietal cortex, average thickness | HC Aβ– | –0.08 [–0.12; –0.04] | 0.02 | –4.04 | 236.49 | 0.06 | < .001*** |
| | HC Aβ + | –0.08 [–0.17; 0.00] | 0.04 | –1.93 | 232.22 | 0.02 | .055 |
| | SCD Aβ + | –0.23 [–0.30; –0.17] | 0.03 | –7.05 | 271.66 | 0.15 | < .001*** |
| | MCI Aβ + | –0.23 [–0.32; –0.15] | 0.04 | –5.29 | 272.78 | 0.09 | < .001*** |
| | DAT Aβ + | –0.60 [–0.72; –0.48] | 0.06 | –9.72 | 302.80 | 0.24 | < .001*** |

[1]Fitted without random slopes due to insufficient variance in random slope estimates.

Abbreviations: AMY, amygdala. BA, Brodmann area. CA, cornu ammonis. CA23DG, cornu ammonis 2, 3, and dentate gyrus. DAT, dementia of the Alzheimer type. ERC, entorhinal cortex. HC, healthy control. IPC, inferior parietal cortex. MCI, mild cognitive impairment. PCC, posterior cingulate cortex. PHC, parahippocampal cortex. PRE, precuneus. RSC, retrosplenial cortex. SCD, subjective cognitive decline. SUB, subiculum. TAIL, hippocampal tail.

**Supplementary Table 6** Pairwise contrasts of diagnostic groups for estimated marginal means of the linear fixed effect of years since baseline on structural MRI markers, derived from linear mixed-effects models.

| Marker | Contrast | *b* [95% C.I.] | *SE* | *t* | *df* | $\eta^2_{partial}$ | *p* |
|---|---|---|---|---|---|---|---|
| Amygdala, volume | HC Aβ+ *versus* HC Aβ – | –0.07 [–0.12; –0.03] | 0.02 | –2.99 | 248.12 | 0.03 | .003** |
| | SCD Aβ + *versus* HC Aβ – | –0.05 [–0.09; –0.01] | 0.02 | –2.69 | 279.61 | 0.03 | .007** |
| | MCI Aβ + *versus* HC Aβ – | –0.11 [–0.16; –0.07] | 0.02 | –5.18 | 298.31 | 0.08 | < .001*** |
| | DAT Aβ + *versus* HC Aβ – | –0.14 [–0.19; –0.08] | 0.03 | –4.89 | 375.01 | 0.06 | < .001*** |
| | SCD Aβ + *versus* HC Aβ + | 0.02 [–0.03; 0.08] | 0.03 | 0.88 | 261.33 | 0.00 | .379 |
| | MCI Aβ + *versus* SCD Aβ + | –0.06 [–0.11; –0.02] | 0.02 | –2.66 | 304.10 | 0.02 | .008** |
| | DAT Aβ + *versus* MCI Aβ + | –0.02 [–0.08; 0.04] | 0.03 | –0.75 | 374.25 | 0.00 | .451 |
| CA1, volume | HC Aβ+ *versus* HC Aβ – | –0.02 [–0.06; 0.02] | 0.02 | –1.11 | 223.84 | 0.01 | .269 |
| | SCD Aβ + *versus* HC Aβ – | –0.02 [–0.05; 0.02] | 0.02 | –0.98 | 246.34 | 0.00 | .326 |
| | MCI Aβ + *versus* HC Aβ – | –0.05 [–0.09; 0.00] | 0.02 | –2.13 | 273.69 | 0.02 | .034* |
| | DAT Aβ + *versus* HC Aβ – | –0.08 [–0.13; –0.02] | 0.03 | –2.62 | 313.32 | 0.02 | .009** |
| | SCD Aβ + *versus* HC Aβ + | 0.01 [–0.04; 0.05] | 0.02 | 0.26 | 237.49 | 0.00 | .795 |
| | MCI Aβ + *versus* SCD Aβ + | –0.03 [–0.08; 0.02] | 0.02 | –1.22 | 277.33 | 0.01 | .225 |
| | DAT Aβ + *versus* MCI Aβ + | –0.03 [–0.10; 0.04] | 0.03 | –0.91 | 309.03 | 0.00 | .365 |
| CA23DG, volume | HC Aβ+ *versus* HC Aβ – | –0.01 [–0.04; 0.03] | 0.02 | –0.28 | 224.97 | 0.00 | .783 |
| | SCD Aβ + *versus* HC Aβ – | –0.03 [–0.06; 0.00] | 0.02 | –1.86 | 247.61 | 0.01 | .064 |
| | MCI Aβ + *versus* HC Aβ – | –0.05 [–0.09; –0.01] | 0.02 | –2.30 | 274.61 | 0.02 | .022* |
| | DAT Aβ + *versus* HC Aβ – | –0.10 [–0.16; –0.04] | 0.03 | –3.49 | 314.54 | 0.04 | < .001*** |
| | SCD Aβ + *versus* HC Aβ + | –0.03 [–0.07; 0.02] | 0.02 | –1.11 | 238.69 | 0.01 | .268 |
| | MCI Aβ + *versus* SCD Aβ + | –0.02 [–0.06; 0.03] | 0.02 | –0.75 | 278.51 | 0.00 | .453 |
| | DAT Aβ + *versus* MCI Aβ + | –0.05 [–0.11; 0.01] | 0.03 | –1.55 | 310.25 | 0.01 | .122 |
| Subiculum, volume | HC Aβ+ *versus* HC Aβ – | –0.02 [–0.06; 0.03] | 0.02 | –0.73 | 227.97 | 0.00 | .463 |
| | SCD Aβ + *versus* HC Aβ – | –0.05 [–0.08; –0.02] | 0.02 | –2.91 | 250.87 | 0.03 | .004** |
| | MCI Aβ + *versus* HC Aβ – | –0.07 [–0.11; –0.03] | 0.02 | –3.13 | 276.79 | 0.03 | .002** |
| | DAT Aβ + *versus* HC Aβ – | –0.12 [–0.17; –0.06] | 0.03 | –3.92 | 316.77 | 0.05 | < .001*** |
| | SCD Aβ + *versus* HC Aβ + | –0.03 [–0.08; 0.01] | 0.02 | –1.46 | 241.86 | 0.01 | .145 |
| | MCI Aβ + *versus* SCD Aβ + | –0.02 [–0.07; 0.03] | 0.02 | –0.75 | 281.32 | 0.00 | .452 |
| | DAT Aβ + *versus* MCI Aβ + | –0.05 [–0.11; 0.02] | 0.03 | –1.38 | 312.60 | 0.01 | .169 |
| Hippocampal tail, volume | HC Aβ+ *versus* HC Aβ – | –0.01 [–0.07; 0.04] | 0.03 | –0.50 | 225.56 | 0.00 | .614 |
| | SCD Aβ + *versus* HC Aβ – | –0.05 [–0.09; 0.00] | 0.02 | –2.12 | 248.28 | 0.02 | .035* |
| | MCI Aβ + *versus* HC Aβ – | –0.03 [–0.09; 0.02] | 0.03 | –1.12 | 275.09 | 0.00 | .263 |
| | DAT Aβ + *versus* HC Aβ – | –0.04 [–0.12; 0.03] | 0.04 | –1.09 | 315.14 | 0.00 | .277 |
| | SCD Aβ + *versus* HC Aβ + | –0.03 [–0.09; 0.03] | 0.03 | –1.09 | 239.33 | 0.00 | .275 |
| | MCI Aβ + *versus* SCD Aβ + | 0.02 [–0.05; 0.08] | 0.03 | 0.48 | 279.13 | 0.00 | .629 |
| | DAT Aβ + *versus* MCI Aβ + | –0.01 [–0.10; 0.08] | 0.04 | –0.22 | 310.86 | 0.00 | .824 |
| Entorhinal cortex, volume | HC Aβ+ *versus* HC Aβ – | –0.07 [–0.12; –0.02] | 0.02 | –3.01 | 225.59 | 0.04 | .003** |
| | SCD Aβ + *versus* HC Aβ – | –0.05 [–0.08; –0.01] | 0.02 | –2.42 | 248.31 | 0.02 | .016* |
| | MCI Aβ + *versus* HC Aβ – | –0.08 [–0.13; –0.03] | 0.02 | –3.29 | 275.10 | 0.04 | .001** |
| | DAT Aβ + *versus* HC Aβ – | –0.17 [–0.24; –0.11] | 0.03 | –5.23 | 315.14 | 0.08 | < .001*** |
| | SCD Aβ + *versus* HC Aβ + | 0.02 [–0.03; 0.08] | 0.03 | 0.90 | 239.35 | 0.00 | .369 |
| | MCI Aβ + *versus* SCD Aβ + | –0.03 [–0.09; 0.02] | 0.03 | –1.25 | 279.14 | 0.01 | .211 |
| | DAT Aβ + *versus* MCI Aβ + | –0.09 [–0.17; –0.02] | 0.04 | –2.42 | 310.86 | 0.02 | .016* |
| BA35, volume | HC Aβ+ *versus* HC Aβ – | –0.07 [–0.12; –0.02] | 0.02 | –2.73 | 229.00 | 0.03 | .007** |
| | SCD Aβ + *versus* HC Aβ – | –0.04 [–0.08; 0.00] | 0.02 | –2.03 | 252.20 | 0.02 | .043* |
| | MCI Aβ + *versus* HC Aβ – | –0.04 [–0.09; 0.01] | 0.03 | –1.68 | 277.97 | 0.01 | .093 |
| | DAT Aβ + *versus* HC Aβ – | –0.16 [–0.22; –0.09] | 0.03 | –4.48 | 317.93 | 0.06 | < .001*** |
| | SCD Aβ + *versus* HC Aβ + | 0.03 [–0.03; 0.08] | 0.03 | 0.94 | 243.11 | 0.00 | .349 |
| | MCI Aβ + *versus* SCD Aβ + | 0.00 [–0.06; 0.05] | 0.03 | –0.08 | 282.71 | 0.00 | .940 |
| | DAT Aβ + *versus* MCI Aβ + | –0.11 [–0.19; –0.03] | 0.04 | –2.80 | 313.81 | 0.02 | .005** |
| BA36, volume | HC Aβ+ *versus* HC Aβ – | –0.03 [–0.06; 0.00] | 0.02 | –1.76 | 224.86 | 0.01 | .080 |
| | SCD Aβ + *versus* HC Aβ – | –0.03 [–0.06; –0.01] | 0.01 | –2.37 | 247.49 | 0.02 | .019* |
| | MCI Aβ + *versus* HC Aβ – | –0.04 [–0.08; –0.01] | 0.02 | –2.50 | 274.53 | 0.02 | .013* |
| | DAT Aβ + *versus* HC Aβ – | –0.18 [–0.23; –0.13] | 0.02 | –7.50 | 314.41 | 0.15 | < .001*** |
| | SCD Aβ + *versus* HC Aβ + | 0.00 [–0.04; 0.03] | 0.02 | –0.17 | 238.58 | 0.00 | .864 |
| | MCI Aβ + *versus* SCD Aβ + | –0.01 [–0.05; 0.03] | 0.02 | –0.58 | 278.40 | 0.00 | .564 |
| | DAT Aβ + *versus* MCI Aβ + | –0.13 [–0.19; –0.08] | 0.03 | –4.90 | 310.12 | 0.07 | < .001*** |
| Parahippocampal cortex, volume[1] | HC Aβ+ *versus* HC Aβ – | 0.03 [–0.02; 0.07] | 0.02 | 1.17 | 218.87 | 0.01 | .243 |
| | SCD Aβ + *versus* HC Aβ – | –0.01 [–0.05; 0.03] | 0.02 | –0.49 | 240.66 | 0.00 | .625 |
| | MCI Aβ + *versus* HC Aβ – | –0.03 [–0.08; 0.01] | 0.02 | –1.44 | 269.19 | 0.01 | .151 |
| | DAT Aβ + *versus* HC Aβ – | –0.14 [–0.21; –0.08] | 0.03 | –4.31 | 304.65 | 0.06 | < .001*** |
| | SCD Aβ + *versus* HC Aβ + | –0.04 [–0.09; 0.01] | 0.03 | –1.39 | 232.34 | 0.01 | .167 |
| | MCI Aβ + *versus* SCD Aβ + | –0.03 [–0.08; 0.03] | 0.03 | –0.95 | 271.62 | 0.00 | .344 |
| | DAT Aβ + *versus* MCI Aβ + | –0.11 [–0.18; –0.03] | 0.04 | –2.82 | 300.99 | 0.03 | .005** |
| Retrosplenial cortex, average thickness | HC Aβ+ *versus* HC Aβ – | 0.01 [–0.04; 0.05] | 0.02 | 0.25 | 223.17 | 0.00 | .804 |
| | SCD Aβ + *versus* HC Aβ – | –0.04 [–0.07; 0.00] | 0.02 | –1.86 | 251.07 | 0.01 | .064 |
| | MCI Aβ + *versus* HC Aβ – | –0.06 [–0.11; –0.02] | 0.02 | –2.67 | 259.02 | 0.03 | .008** |
| | DAT Aβ + *versus* HC Aβ – | –0.16 [–0.22; –0.09] | 0.03 | –4.89 | 284.46 | 0.08 | < .001*** |
| | SCD Aβ + *versus* HC Aβ + | –0.04 [–0.09; 0.01] | 0.03 | –1.58 | 238.07 | 0.01 | .115 |
| | MCI Aβ + *versus* SCD Aβ + | –0.03 [–0.08; 0.02] | 0.03 | –1.06 | 264.23 | 0.00 | .289 |
| | DAT Aβ + *versus* MCI Aβ + | –0.09 [–0.17; –0.02] | 0.04 | –2.54 | 280.32 | 0.02 | .012* |
| Posterior cingulate cortex, average thickness | HC Aβ+ *versus* HC Aβ – | 0.02 [–0.05; 0.09] | 0.04 | 0.52 | 718.32 | 0.00 | .601 |
| | SCD Aβ + *versus* HC Aβ – | –0.02 [–0.08; 0.04] | 0.03 | –0.69 | 721.12 | 0.00 | .490 |
| | MCI Aβ + *versus* HC Aβ – | –0.05 [–0.13; 0.02] | 0.04 | –1.40 | 721.37 | 0.00 | .162 |
| | DAT Aβ + *versus* HC Aβ – | –0.25 [–0.35; –0.15] | 0.05 | –4.92 | 723.85 | 0.03 | < .001*** |
| | SCD Aβ + *versus* HC Aβ + | –0.04 [–0.12; 0.04] | 0.04 | –0.97 | 720.35 | 0.00 | .334 |

| | | | | | | | |
|---|---|---|---|---|---|---|---|
| | MCI Aβ + *versus* SCD Aβ + | –0.03 [–0.11; 0.05] | 0.04 | –0.76 | 722.51 | 0.00 | .445 |
| | DAT Aβ + *versus* MCI Aβ + | –0.20 [–0.31; –0.08] | 0.06 | –3.39 | 723.55 | 0.02 | < .001*** |
| Precuneus, average thickness | HC Aβ+ *versus* HC Aβ – | –0.02 [–0.10; 0.06] | 0.04 | –0.48 | 230.59 | 0.00 | .632 |
| | SCD Aβ + *versus* HC Aβ – | –0.10 [–0.17; –0.04] | 0.03 | –3.10 | 258.21 | 0.04 | .002** |
| | MCI Aβ + *versus* HC Aβ – | –0.11 [–0.20; –0.03] | 0.04 | –2.77 | 266.38 | 0.03 | .006** |
| | DAT Aβ + *versus* HC Aβ – | –0.32 [–0.43; –0.22] | 0.06 | –5.88 | 294.35 | 0.11 | < .001*** |
| | SCD Aβ + *versus* HC Aβ + | –0.08 [–0.17; 0.01] | 0.05 | –1.83 | 245.29 | 0.01 | .069 |
| | MCI Aβ + *versus* SCD Aβ + | –0.01 [–0.10; 0.08] | 0.05 | –0.27 | 272.25 | 0.00 | .788 |
| | DAT Aβ + *versus* MCI Aβ + | –0.21 [–0.33; –0.09] | 0.06 | –3.32 | 289.78 | 0.04 | .001** |
| Inferior parietal cortex, average thickness | HC Aβ+ *versus* HC Aβ – | 0.00 [–0.10; 0.09] | 0.05 | –0.10 | 231.83 | 0.00 | .920 |
| | SCD Aβ + *versus* HC Aβ – | –0.15 [–0.23; –0.08] | 0.04 | –3.89 | 259.20 | 0.06 | < .001*** |
| | MCI Aβ + *versus* HC Aβ – | –0.16 [–0.25; –0.06] | 0.05 | –3.17 | 267.36 | 0.04 | .002** |
| | DAT Aβ + *versus* HC Aβ – | –0.52 [–0.65; –0.39] | 0.07 | –7.99 | 295.33 | 0.18 | < .001*** |
| | SCD Aβ + *versus* HC Aβ + | –0.15 [–0.25; –0.04] | 0.05 | –2.74 | 246.43 | 0.03 | .007** |
| | MCI Aβ + *versus* SCD Aβ + | 0.00 [–0.11; 0.10] | 0.05 | –0.05 | 273.29 | 0.00 | .959 |
| | DAT Aβ + *versus* MCI Aβ + | –0.37 [–0.51; –0.22] | 0.08 | –4.89 | 290.78 | 0.08 | < .001*** |

[1]Fitted without random slopes due to insufficient variance in random slope estimates.

Abbreviations: AMY, amygdala. BA, Brodmann area. CA, cornu ammonis. CA23DG, cornu ammonis 2, 3, and dentate gyrus. DAT, dementia of the Alzheimer type. ERC, entorhinal cortex. HC, healthy control. IPC, inferior parietal cortex. MCI, mild cognitive impairment. PCC, posterior cingulate cortex. PHC, parahippocampal cortex. PRE, precuneus. RSC, retrosplenial cortex. SCD, subjective cognitive decline. SUB, subiculum. TAIL, hippocampal tail.
